# Identification of Spatial Proteomic Signatures of Colon Tumor Metastasis: A Digital Spatial Profiling Approach

**DOI:** 10.1101/2022.12.04.22283073

**Authors:** Joshua J. Levy, John P. Zavras, Eren M. Veziroglu, Mustafa Nasir-Moin, Fred W. Kolling, Brock C. Christensen, Lucas A. Salas, Rachael E. Barney, Scott M. Palisoul, Bing Ren, Xiaoying Liu, Darcy A. Kerr, Kelli B. Pointer, Gregory J. Tsongalis, Louis J. Vaickus

## Abstract

Over 150,000 Americans are diagnosed with colorectal cancer (CRC) every year, and annually over 50,000 individuals will die from CRC, necessitating improvements in screening, prognostication, disease management, and therapeutic options. CRC tumors are removed en bloc with surrounding vasculature and lymphatics. Examination of regional lymph nodes at the time of surgical resection is essential for prognostication. Developing alternative approaches to indirectly assess recurrence risk would have utility in cases where lymph node yield is incomplete or inadequate. Spatially dependent, immune cell-specific (e.g., Tumor Infiltrating Lymphocytes– TILs), proteomic, and transcriptomic expression patterns inside and around the tumor - the tumor immune microenvironment (TIME) - can predict nodal/distant metastasis and probe the coordinated immune response from the primary tumor site. The comprehensive characterization of TILs and other immune infiltrates is possible using highly multiplexed spatial omics technologies, such as the GeoMX Digital Spatial Profiler (DSP). In this study, machine learning and differential co-expression analyses helped identify biomarkers from DSP-assayed protein expression patterns inside, at the invasive margin, and away from the tumor, associated with extracellular matrix remodeling (e.g., GZMB, fibronectin), immune suppression (e.g., FOXP3), exhaustion and cytotoxicity (e.g., CD8), PD-L1 expressing dendritic cells, neutrophil proliferation, amongst other concomitant alterations. Further investigation of these biomarkers may reveal independent risk factors of CRC metastasis that can be formulated into low-cost, widely available assays.

## Introduction

Colorectal cancer (CRC) has the fourth highest cancer incidence rate in the United States, and carries a lifetime risk of roughly 4%. The incidence of colorectal cancer is correlated with increasing age with the majority of colorectal cancer cases occurring in patients over the age of 50. However, in the United States the incidence of colorectal cancer in patients under the age of 50 is rising. The reason for this sudden increase is not known, as the disease etiology is thought to be a complex interaction of dietary patterns, environmental exposures, and genetic influences. Additionally, CRC incidence varies widely around the world, with the highest rates reported in Australia, New Zealand, Europe, and North America further supporting the complexity of the disease. According to the National Cancer Institute’s Surveillance Epidemiology and End Results program (SEER), disparities exist by age, sex, and ethnicity in disease incidence nationwide. Despite varying incidence rates, CRC is the third largest contributor to cancer deaths worldwide, indicating a significant unmet need to improve curative-intent therapy for adequately identifying and treating CRC to prevent death ^1–3^.

Early-stage colorectal cancer that has not spread to lymph nodes or other distant sites has a 5-year survival rate of 91%; however, if it has spread to the regional lymph nodes, the 5-year survival rate drops to 72%, an important factor in patient outcomes. Pathologic Tumor, Node, Metastasis (pTNM) stage at presentation is considered the most important factor for ascertaining CRC prognosis. CRC staging elements include: the level of tumor invasion (T0-4b), regional lymph node metastasis (N0-2b), and distant metastasis (M0-1). Distant metastasis at diagnosis is associated with a 5-year survival rate of only 14%. Approximately 37% of patients are diagnosed with localized disease, 36% are diagnosed with regional lymph node spread, and 22% with distant metastasis. Identification of regional nodal disease often indicates the usage of adjuvant therapies (e.g., chemotherapy), as regional nodal positivity is a risk factor for tumor recurrence ^4,5^.

Processes governing cellular migration and metastasis involve cellular biochemical alterations related with primary tumor formation. Through rapid mitotic activity and the accumulation of genomic instability, cells gain the capacity to invade mesenchymal tissue and vasculature. In circulation, tumor cells migrate through the intravascular and extravascular systems, evade DNA damage response pathways, prime sites for metastasis, and establish a favorable micro-environment for metastasis including activation of T regulatory cells and angiogenesis. Outside of the standard pTNM staging, some hypothesize that tumor border morphology is a strong, independent prognostic factor (e.g., tumor budding). Other specific pathologic findings of poor prognosis include a poorly defined border, invasion through mucosal layers without expected stromal reaction, and focal de-differentiation. Metastatic potential has also been shown to vary by primary tumor site (e.g., left-sided) ^6^. These findings may be specific to common progression pathways (e.g., APC, MSI– microsatellite instability, CIMP– CpG Island Methylator Phenotype).

Several studies concur on the importance of the tumor immune microenvironment (TIME) in CRC prognosis. Specifically, high densities of tumor-infiltrating lymphocytes (TIL) have been shown to improve prognosis and the 5-year survival rate of patients with CRC metastases. In addition to spatial distribution and density, previous studies have shown that the type, activation state, and location of infiltrating lymphocytes determine the tumor microenvironment’s immune response and its antitumoral effectiveness. In general, the presence of TILs confers a favorable prognosis. TIL cells appear to have more potent and directed anti-tumor effects than peripheral blood circulating lymphocytes ^7–9^. Various tumor-specific characteristics, including mismatch repair alterations, determine TIL’s effect on the tumor microenvironment (TME) and prognosis ^10–12^. While ample evidence suggests that while the overall presence of immune infiltration carries a favorable prognosis, not all immune lineages support these favorable findings.

Current methods of prognosticating recurrence and survival are somewhat crude and can be improved. Specimen inadequacy and inadequate lymph nodes yield are important limitations to current prognostication methods ^13–17^. This incomplete or inadequate assessment can affect the accuracy of tumor staging and subsequent disease management options, such as whether a patient should receive adjuvant chemotherapy. Studies have also demonstrated that patients who receive extended lymphadenectomies have better outcomes; however, this is not the standard of care and can cause increased morbidity. Multiplexed genomic, proteomic, and transcriptomic assays of tumors have revealed an incredible variation both at the level of the host, the tumor, and the tumor’s micro-environment, along with complex regulatory networks and interactions. However, most of these findings are still in the discovery phase and have not been validated clinically, with the “weak link” of such multiplexed methods being the origin of the examined cells. New evidence strongly suggests that cells’ origin and anatomical location play a significant role in prognostication. Multiplexed spatial molecular cancer tools have recently been developed, creating a new frontier in cancer diagnosis, treatment, and prognosis. In CRC specifically, a lot remains to be learned, such as information on cell-type specific molecular alterations (e.g., transcriptome expression) within unique spatial arrangements related to colon cancer metastasis ^18^. The development of spatial omics technologies such as 10x Genomics Spatial Transcriptomics (ST) or GeoMX Digital Spatial Profiling (DSP) has enabled multiplexing findings (e.g., whole transcriptome, WTA) at incredible spatial resolution^28^. The GeoMX platform first deposits an RNA/Protein barcode for expression profiling across an entire tissue slide. Fluorescent antibodies that highlight various tissue types stain the slide to highlight relevant structures for the selection of Regions of Interest (ROI). Within selected regions of interest (which can vary from single-cell sized to nearly a centimeter squared), ultraviolet light is used to cleave barcodes from substrate selectively, and these barcodes are retrieved for quantitation using nCounter and NGS sequencing. Analysis of such data may reveal spatially variable gene expression, characteristic spatial patterns of expression (which may inform spatially variable cell type proportions), or information about how cells communicate to elicit an immune response.

As an application of these techniques, GeoMX platform has been used to study the differences between TILs and non-tumor/stromal lymphocytes in tumors driven by microsatellite instability ^19^. This study uncovered expression patterns in intratumoral T-cells and extratumoral T-cells related to cytolytic activity and cell-cell interactions. Another study found a correlation between spatial statistics from 55 fluorescently tagged antibodies and the 5-year risk of CRC progression ^20,21^. Other studies include the role of neutrophils in poor prognosis ^22^ and how the spatial distance between key TIL lineages may be influenced by PD-L1 expression ^20,21^. Finally, the spatial relationship between lymphocytes and tumor budding has been shown to be a prognostic predictor ^23,24^. Few studies have attempted to use information on tissue context specific immune cell expression to predict the metastastic potential of CRC tumors.

Several factors complicate assessments for the presence and prevention/inhibition of tumor metastasis, chiefly considering the capacity to surgically resect the positive lymph nodes as well as establish etiological models of metastasis that are targetable through commensurate emerging therapeutics (e.g., immunotherapies). For instance, an incomplete lymph node dissection can potentially result in a false negative finding, inaccurately determining recurrence risk. These deficiencies necessitate the development of tools to reduce the potential for inadequate assessments or to fill in the missing information. In this study, we aim to characterize spatial immunological correlates of nodal and distant metastasis from the primary tumor site through the application of digital spatial profiling.

## Methods

### Methods Overview

Figure 1. shows a graphical overview of our methodology. In brief, we: 1) collected 36 resected colorectal tumor specimens; 2) using a spatial proteomics assay, profiled immune cells within three spatial architectures; and 3) identified biomarkers/lineages associated with nodal and distant metastasis within these architectures, controlling for local invasiveness. Biomarkers were inferred through differential expression analyses, and we assessed the potential for two biomarkers at a time to carry additional predictive value through the following assessments: 1) relative expression between the biomarkers (i.e.,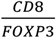;2) interactions (i.e., conditional on one cell-type, what is the association of another and metastasis); and 3) differential coexpression.

**Figure 1:**
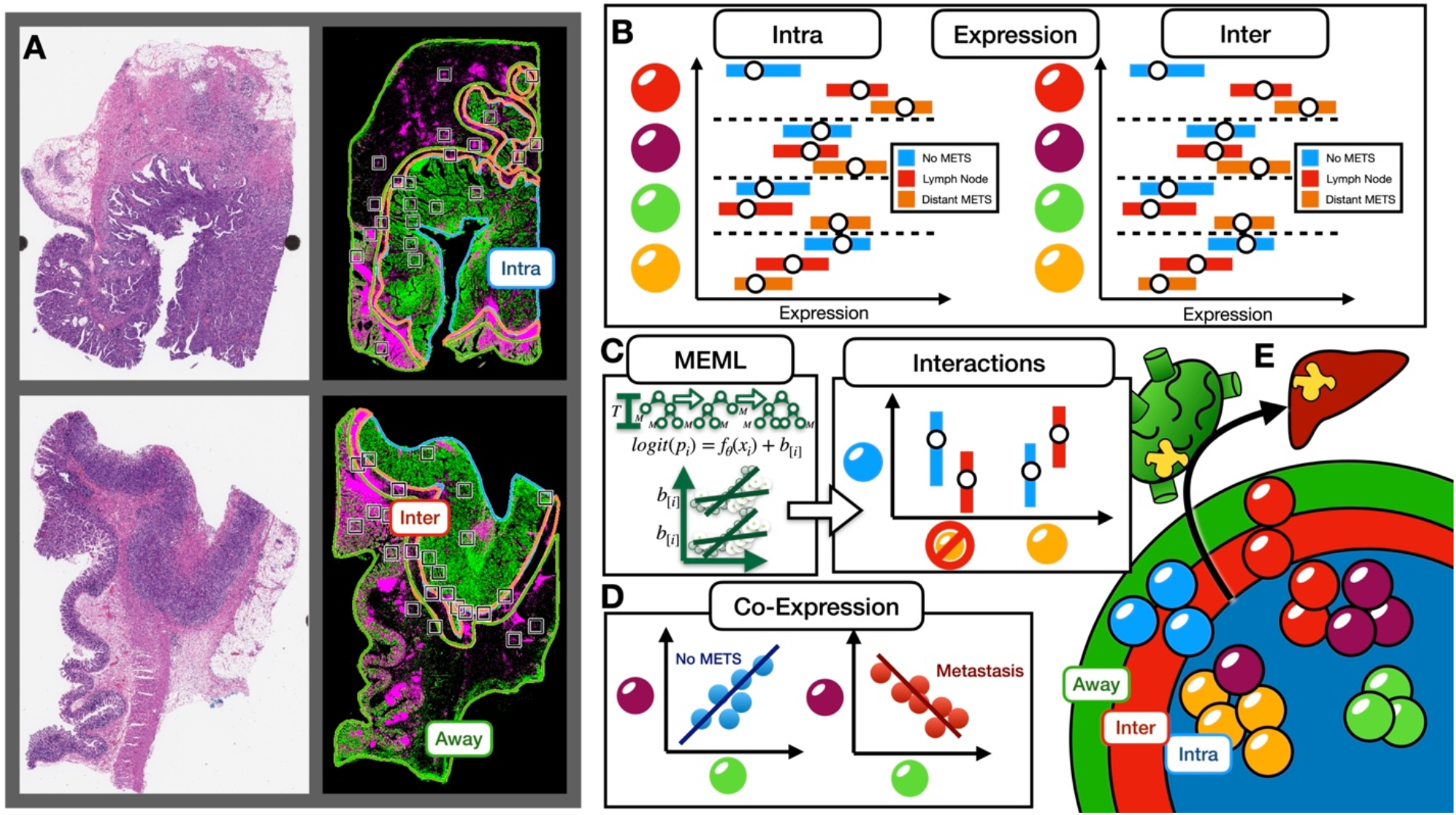
Study Overview: A) H&E and IF stained slide used to help place ROI for profiling within three distinct architectures: *intratumoral, interface, away*; B) within distinct architectures, expression of specific lineages/protein markers predictive of metastasis status; C) Mixed-effects machine learning models uncover statistical interactions between specific immune cell types (i.e., different risk of metastasis for specific cell type conditional on another cell type); D) Differential coexpression patterns identify correlations between markers that are metastasis specific; E) Identifying predictive biomarkers for nodal and distant metastasis within these regions

### Data Acquisition and Preprocessing

Thirty-six colon adenocarcinoma resections performed at Dartmouth Hitchcock Medical Center (DHMC) from 2016 to 2019 were selected for digital spatial profiling with IRB approval. Approximately half of them showed local invasion but no nodal or distant metastasis (no METS), and the other half showed nodal and/or distant metastasis (METS). Of the cases with concurrent metastasis, all cases exhibited local lymph node involvement– about half of these cases metastasized to distant sites. Sample size was determined based on feasibility of this pilot study and through an empirical power analysis which simulated data from the statistical models introduced in subsequent sections. The cohort was restricted to stage pT3 assignments under the pTNM staging system, which balances the impacts of local invasion, nodal and distant metastasis for prognostication. T-stage refers to the degree of invasion at the local site. By restricting the T stage, we sought to identify markers that provide prognostic value beyond that offered through the current prognostic staging system. Cases were matched between the non-metastatic and metastatic groups based on tissue size (measured through connected component analysis of whole slide images), tumor grade, mismatch repair (MMR) dysregulation status (*dMMR*– deficiency, *pMMR*– proficient; as assessed through IHC), site of the tumor (e.g., left or right colon), age, and sex. Matching and randomization was achieved by conducting Fisher’s exact tests and two-sample t-tests after iterative resampling (**Table 1**). MMR deficiencies (*dMMR*) reflect the loss of staining in at least one of four mismatch repair genes (MLH1, PMS2, MSH2, MSH6). As MSH2 and MSH6 alterations were relatively rare for cases within the queried time periods, *dMMR* status was reported from alterations to either MLH1 or PMS2 (MSH2 and MSH6 alterations were not present in this cohort) ^25,26^.

**Table 1:** Patient Cohort Characteristics; stratified by metastasis status (yes/no). For patients with metastasis, stratified by whether there was only nodal involvement or both nodal and distant involvement

|  | No<br>Metastasis | Metastasis | p-value | Lymph<br>Node<br>Only | Distant +<br>Lymph<br>Node | p-value |
| --- | --- | --- | --- | --- | --- | --- |
| **N** | 16 | 19 |  | 8 | 11 |  |
| **pMMR (%)** | 10 (62.5) | 12 (63.2) | 1 | 6 (75.0) | 6 (54.5) | 0.667 |
| **grade (%)** |  |  | 0.789 |  |  | 0.408 |
| **1** | 8 (50.0) | 11 (57.9) |  | 6 (75.0) | 5 (45.5) |  |
| **2** | 4 (25.0) | 3 (15.8) |  | 1 (12.5) | 2 (18.2) |  |
| **3** | 4 (25.0) | 5 (26.3) |  | 1 (12.5) | 4 (36.4) |  |
| **site (%)** |  |  | 0.936 |  |  | 0.142 |
| **cecum** | 6 (37.5) | 5 (26.3) |  | 1 (12.5) | 4 (36.4) |  |
| **hepatic flexure** | 1 (6.2) | 1 (5.3) |  | 0 (0.0) | 1 (9.1) |  |
| **left colon** | 2 (12.5) | 2 (10.5) |  | 1 (12.5) | 1 (9.1) |  |
| **rectum** | 1 (6.2) | 3 (15.8) |  | 3 (37.5) | 0 (0.0) |  |
| **right colon** | 2 (12.5) | 3 (15.8) |  | 2 (25.0) | 1 (9.1) |  |
| **sigmoid** | 1 (6.2) | 3 (15.8) |  | 0 (0.0) | 3 (27.3) |  |
| **splenic flexure** | 1 (6.2) | 1 (5.3) |  | 1 (12.5) | 0 (0.0) |  |
| **transverse colon** | 2 (12.5) | 1 (5.3) |  | 0 (0.0) | 1 (9.1) |  |
| **sex = M (%)** | 9 (56.2) | 11 (57.9) | 1 | 6 (75.0) | 5 (45.5) | 0.414 |
| **age (mean (SD))** | 71.06 (12.87) | 66.21 (16.29) | 0.342 | 67.75<br>(17.53) | 65.09<br>(16.10) | 0.736 |
| **N-Stage (%)** |  |  |  | n/a |  | 0.074 |
| **0** | 16 (100.0) | 0 (0.0) |  | 0 (0.0) | 0 (0.0) |  |
| **1a** | 0 (0.0) | 9 (47.4) |  | 2 (25.0) | 7 (63.6) |  |
| **1b** | 0 (0.0) | 4 (21.1) |  | 2 (25.0) | 2 (18.2) |  |
| **1c** | 0 (0.0) | 2 (10.5) |  | 0 (0.0) | 2 (18.2) |  |
| **2a** | 0 (0.0) | 2 (10.5) |  | 2 (25.0) | 0 (0.0) |  |
| **2b** | 0 (0.0) | 2 (10.5) |  | 2 (25.0) | 0 (0.0) |  |

Tissue blocks were sectioned with 5-µm thickness and stained with fluorescent-labeled antibodies (highlighting tumor (PanCk), immune cells (CD45), and nuclei (SYTO13)). Fluorescent antibodies were covalently linked to photocleavable oligonucleotide tags associated with a targeted panel of Immune Cell Profiling and Tumor Immune Environment protein markers (40 total markers). Sections were visualized using the GeoMX digital spatial profiling (DSP) instrument, which displayed immunofluorescent (IF) images. Subsequent sections were stained with hematoxylin and eosin (H&E) and scanned using the Leica Aperio-AT2 scanner at 20x. H&E images were stored in SVS format (8-bit color channels). IF whole slide images (WSI) were stored in TIFF format (16-bit unsigned color channels; one channel per stain).

A GI pathologist used the ASAP annotation software (https://computationalpathologygroup.github.io/ASAP/) to view the H&E and IF whole slide images (WSI) side by side to annotate sections and spatially resolved immune populations based on three distinct macroarchitectural regions: intratumoral (*intra*), tumor-immune interface (*inter*) and away from the tumor (*away*) ^27^. These regions were outlined using polygonal/spline annotations. Eight regions of interest (ROI; square grids of maximal spatial dimensions allowable by the GeoMx DSP instrument) were placed within each annotated region per slide (24 ROI per slide) using a semi-autonomous placement system. The ROI were uploaded and registered to the DSP IF images. An automated process initially selected a random distribution of 8 potential ROI per annotated region. When ROI were suboptimal (e.g., insufficient CD45 staining, incorrect region; determined through a visual assessment), pathologists manually adjusted to the nearest appropriate region. Immune cells were isolated within each ROI via image segmentation of the CD45 stain (to establish pixel-wise locations with CD45+SYTO13+PanCk-staining), followed by a connected component analysis. Segmented ROI were profiled through targeted ultraviolet (UV) cleavage of attached oligo tags. The Nanostring nCounter was used to quantify immune cell protein expression across 40 immuno-oncology markers. Four slides were profiled per DSP batch. Tissue lifting after cover-slipping procedures led to additional case inclusion/exclusion. Due to issues with de-coverslipping one of the tissue slides (e.g., tear), one sample was removed from the set (n=35). Returned data included protein expression measurements for each ROI, tagged with positional x and y coordinates, an ROI-specific nuclei count, and co-registered H&E and IF slides from the same section. ROIs were filtered based on expression relative to the negative control (825 ROIs remaining after removal of one case and 15 additional ROIs), normalized to IgG isotype controls (Rb-IgG, Ms-IgG1; Ms IgG2a demonstrated significant batch effects) across batches after comparison to other normalization methods (ERCC, nuclei count/area, housekeepers), and log2 transformed. ROI were further labeled with mismatch repair deficiency status (using MLH1/PMS2 deficiency as assessed through immunohistochemistry-IHC, as a proxy), age and sex, site of origin/metastasis, tumor grade, nodal and distant metastasis status, and macro-architectural region (intra, inter, away).

#### Differential Expression to Establish Clinical Markers of Metastasis

The following bayesian hierarchical linear regression models were fit to predict log2-transformed protein expression to establish associations with metastasis (*mets* used to separately indicate nodal metastasis, distant metastasis, nodal or distant metastasis): 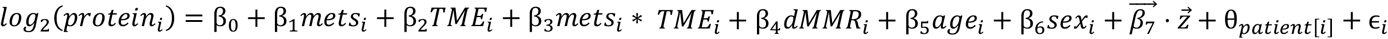. An interaction term between *mets* and *TME* allowed for the evaluation of metastasis conditioned on the macroarchitecture (*TME* ∈ {*intra, inter, away*}), adjusting for potential confounding (*dMMR*, age, sex). Batch and case-level variation were captured with random intercepts, θ. Residual technical variation from the isotype controls was modeled using 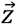. Effect estimates were communicated using the median posterior sample of the effect estimate, 95% high-density posterior credible interval (similar to the confidence interval), posterior probability of direction (*pd*), transformed into a value correlated to the p-value (*p* ≈ 2 ∗ (1 − *pd*)) to communicate the significance of the effect, with *post hoc* comparisons via estimated marginal means through the *emmeans* software package (R statistical programming language v4.1) to report metastasis-related markers by macroarchitecture^28,29^. A weakly informative prior centered around 0 was chosen as type I error control in favor of a multiple comparisons adjustment given the nature of this exploratory analysis though we emphasize the exploratory nature of this assessment ^30,31^.

These associations were also reported separately for patients with/without MSI via the following statistical model:

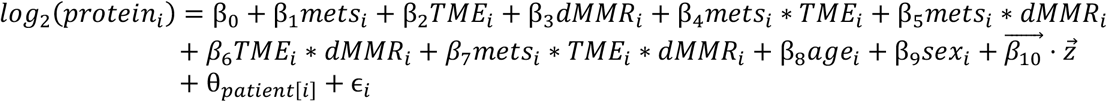

The three-way interaction between metastasis, MSI status, and TME architecture was interrogated using *emmeans* to report metastasis-related markers, conditional on macroarchitecture and MSI status.

We displayed the effect estimates for each analysis for each protein and their corresponding p-values using volcano plots. Expression for markers with large effect sizes and significant p-values (given an alpha significance level of 0.05) were visualized using boxplots. Statistical analyses were performed using the *brms* package from the *R v4*.*1* statistical programming language, which leverages Hamiltonian Monte Carlo (HMC) techniques in *Stan* to compute bayesian effect estimates ^32–34^. HMC models were run using the Discovery computing cluster at Dartmouth College.

#### Relative Expression between Proteins as Additional Markers of Metastasis

In addition to evaluating specific cellular subsets independently, the relative abundance of different immune cell subsets could also point to metastasis-related factors. We modeled the relative expression between two protein markers using the following statistical models:

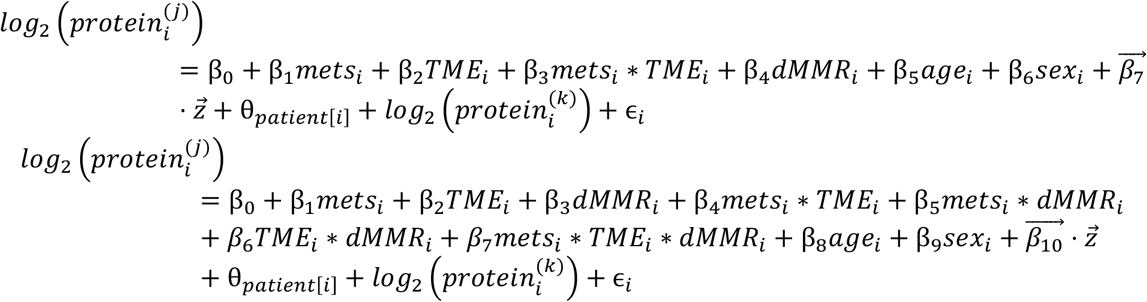

Where the offset term, 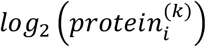 is used to model the relative abundances between immune cell lineages *j* and *k*. Similar posthoc comparisons and displays (i.e., volcano plots, boxplots) were constructed based on the relative proportion between marker expression: 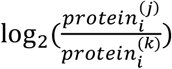

#### Machine Learning Classifiers to Report Salient Effect Modifiers

We developed a set of classifiers to estimate the probability of tumor metastasis (lymph node, distant, any) based on all markers (*x*_*i*_) within distinct architectures (*intra, inter, away*) by fitting nine tree-boosting models, 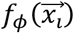, in a Mixed Effects Machine Learning modeling framework (MEML), which leveraged Gaussian Process Boosting (GPBoost) decision trees^35,36^: 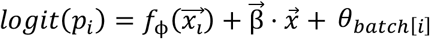. Salient cross-level interactions were identified from the MEML method using the *interactiontransformer* package in Python v3.7, which scores interactions using SHAP and selects the top interactions using a knee locator method. Interactions were applied to a Bayesian hierarchical logistic regression model to report pertinent effect modifiers (e.g., effect of CD20, conditional on age; interactions encapsulated in 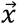)^27,37^: 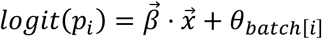. As many interactions were initially selected, features were selected using the Horseshoe LASSO method and the remaining features were fit with weakly informative priors^38,39^. Effect estimates for salient effect modifiers were reported similar to the previous sections. Boxplots were generated for select interactions.

#### Differential Coexpression Networks

We constructed differential coexpression networks to identify sets of genes whose co-expression differed by metastasis status. Coexpression between all pairs of genes was assessed using repeated measure correlation (via the *rmcorr* R package). We adapted the CDS framework for this analysis, which evaluates whether log2-transformed protein coexpression was: Conserved (C), Specific (S), or Differential (D) between patient cohorts with and without metastasis ^40,41^. Each pair of proteins were scored for each of these criteria, which was used to generate networks that pointed to co-expressed proteins that were metastasis-related. A total of nine networks were constructed for combinations of outcomes (lymph node metastasis, distant, any) and distinct architectures (*intra, inter, away*). Important markers for each network were identified through the calculation of the eigenvector centrality on a weighted adjacency matrix.

#### Web Application for Result Viewing

As we discuss only a small subset of comparisons made for this study, we developed an RShiny web application for viewing the study findings ^42^. This application features the ability to view volcano plots, boxplots, differential coexpression, and classifier results from all study comparisons and all numerical findings, including those featured outside of the present work. The Rshiny application is available at the following URL: https://levylab.shinyapps.io/ViewColonDSPResults (**user**: edit_user, **password**: cgat2022).

## Results

This section reports metastasis associations identified using the Digital Spatial Profiler for individual tissue architectures (*intra, inter, away*). First, we sought to establish individual protein markers which correlated with tumor metastasis. Then, we sought to report disease associations that existed by assessing the following markers in tandem: 1) relative abundance/expression, 2) protein interactions, and 3) differential coexpression.

### Report of Clinical Characteristics for Cohort

In **Table 1**, we have included patient demographic characteristics, stratified by: a) metastasis status and b) restricted to patients with metastases, whether the involvement was local or distant. Results indicate that the cohort is well-matched based on MMR status, grade, primary site, sex, age, and N-stage.

### Differential Expression Results

#### Intratumoral

First, we sought to identify metastasis-related markers within distinct macroarchitectures. In general, FOXP3 and CD66b exhibited decreased expression inside the tumor for patients with metastasis (**Supplementary Figures 1-2**). When restricted to MMR-deficient patients, PD-L2 expression was negatively associated with metastasis in the intratumoral region. CD8 was positively associated with lymph node metastasis in the intratumoral regions (**Figure 2**), regardless of MMR status, while CD44 expression was positively associated with distant metastasis. For MMR-deficient patients, GZMB, PD-L1, and Beta-2-Microglobulin were associated with distant metastasis (**Supplementary Figure 3**).

**Figure 2:**
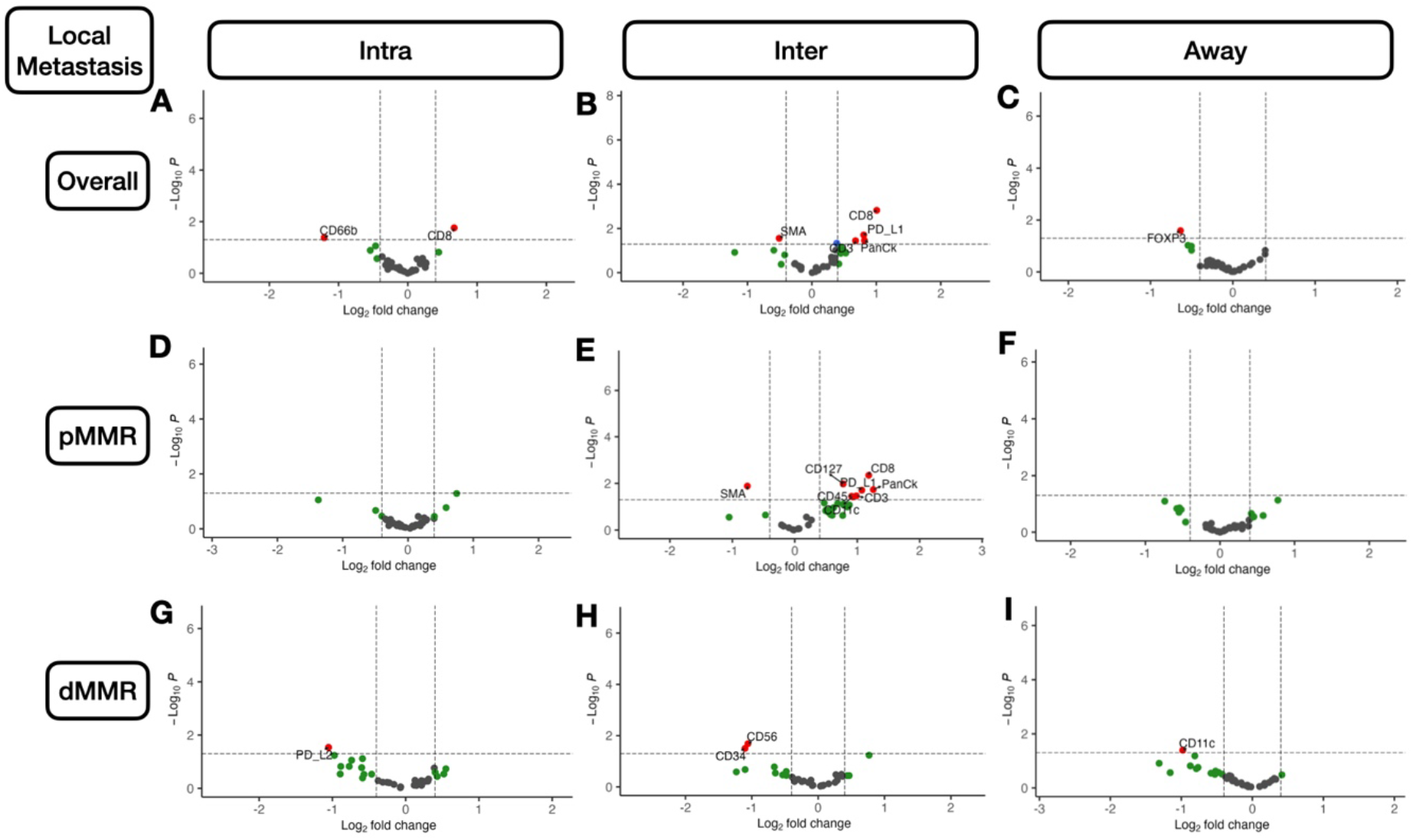
Differentially Expressed Protein Markers of Local Metastasis: A-I) Results stratified by tissue architecture: A,D,G) *Intratumoral*; B,E,H) *peritumoral*; C,F,I) *away/stroma*; results also stratified by MMR-status: D-F) *pMMR*; G-I) *dMMR*; statistical significance cutoff at α = 0.05; x-axis indicates effect size and directionality (positive x-value indicates metastasis-related marker; negative indicates decreased metastasis risk); y-axis indicates effect significance (positive y-value indicates lower p-value)

#### Interface

In the tumor-immune invasive interface, increased expression of GZMB was associated with any metastasis. For patients with MMR deficiency, CD56 expression was negatively correlated with metastasis. Meanwhile, CD8, PD-L1, and CD3 were positively associated with lymph node metastasis (**Figure 3**). In addition to these markers, CD127, CD3, and CD11c were also positively associated with lymph node metastasis for microsatellite-stable tumors. For MMR-deficient patients, CD34 and CD56 expression were reduced for patients with lymph node metastasis. The presence of GZMB in patients with MMR deficiency was positively associated with distant metastasis.

**Figure 3:**
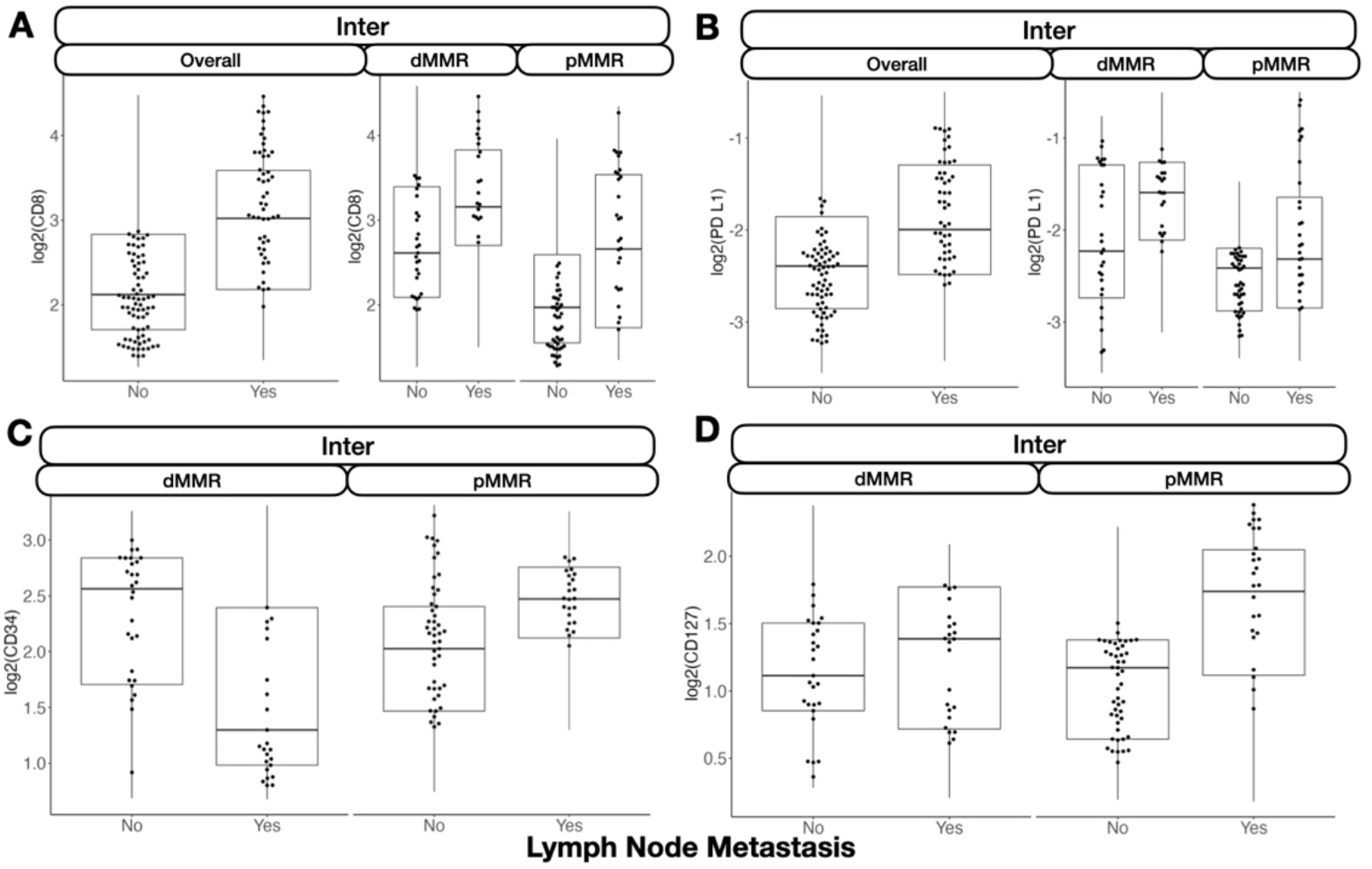
Select protein marker expression for biomarkers predictive of nodal metastasis, stratified by MMR-status: A) CD8 at the *interface*; B) PD-L1 at the *interface*; C) CD34 at the *interface*; D) CD127 at the *interface*. Marker expression plotted in beeswarm plots were filtered based on the detection of outliers using a modified Tukey outlier test– after this initial filtering, only points between the 10% and 90% quantiles for each stratum were included

#### Away

Away from the tumor, several factors were associated with metastasis. In general, FOXP3 and CD14 expression were negatively associated with metastasis. While CD14 expression was especially relevant for patients without microsatellite instability, FOXP3 was salient for MMR-deficient patients. For these patients, CD11c was associated with no tumor metastasis. FOXP3 and CD11c were associated with no lymph node metastasis for patients regardless of MMR status and MMR-deficient patients, respectively. CD11c, Ki-67, and FOXP3 were associated with no distant metastasis, and GZMB expression was associated with distant metastasis for patients with MMR deficiency (**Supplementary Figure 4**).

A complete listing of differentially expressed markers and the relevant statistical findings can be found in the Shiny application.

### Relative Expression Results

#### Nodal Metastasis

We found that the relative abundance between specific immune cell lineages waswashighly predictive of metastasis. For instance, the ratio between CD66b and CD8 expression in the intratumoral region was negatively associated with lymph node metastasis (**Figure 4**). This trend was similar for the ratio between FOXP3 and PD-L1, but only for microsatellite-stable patients. The relative expressions between CD8/CTLA4, and CD8/CD56 were associated with lymph node metastasis at the tumor interface.

**Figure 4:**
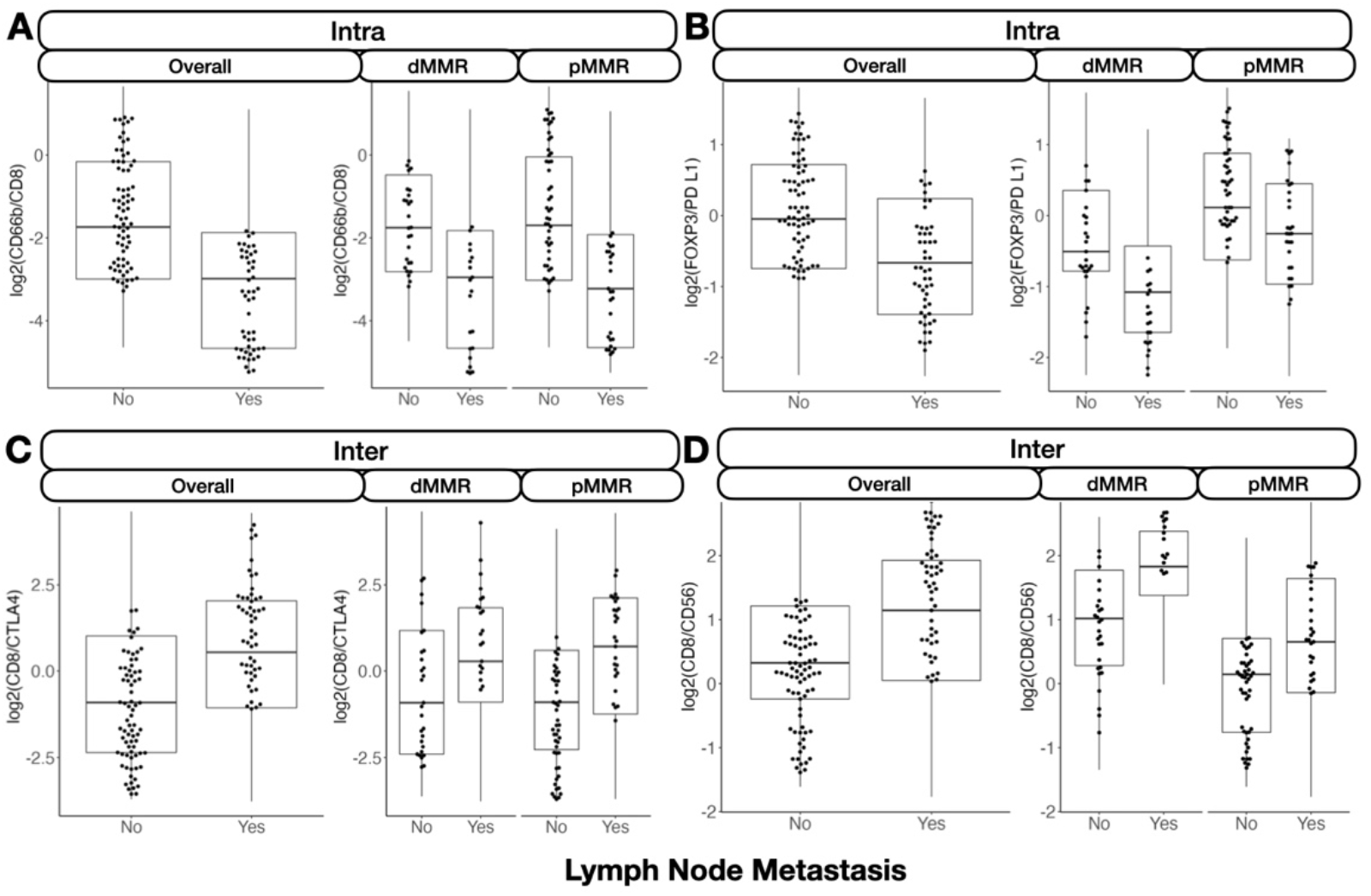
Relative protein expression between markers predictive of nodal metastasis, stratified by MMR-status: A) CD66b/CD8 inside the tumor; B) FOXP3/PD-L1 inside the tumor; C) CD8/CTLA4 at the *interface*; D) CD8/CD56 at the *interface*. Marker expression plotted in beeswarm plots were filtered based on the detection of outliers using a modified Tukey outlier test– after this initial filtering, only points between the 10% and 90% quantiles for each stratum were included.

#### Distant Metastasis

For distant metastasis, the relative expression between CD8 and CD4 as compared to PD-L1 inside the tumor was negatively associated with tumor metastasis for microsatellite-stable patients (**Supplementary Figures 5-6**). At the interface, the relative expression between CD11c and GZMB was associated with no metastasis, while away from the tumor, CD11c as compared to CD34 were heavily negatively associated with distant metastasis.

A complete listing of relative abundance differences can be found in the Shiny application.

### Salient Effect Modifiers

#### Overall Metastasis

Several protein interactions were identified using the classifiers. Of interest, immune cells in the intratumoral region expressing the PD-L1 surface antigen were at reduced risk of metastasis with higher CD34 expression (marker for immune cell stem-ness and vascular endothelial cells). Cells lacking this antigen demonstrated an increased risk of metastasis with higher CD34 expression. Similar relationships were noted between fibronectin and CD27 at the tumor interface– CD27+ cells demonstrated increased metastasis risk as compared to CD27-cells for low levels of fibronectin; the opposite relationship was noted for fibronectin + cells. Away from the tumor, TNF receptor family member cells (CD40+) which also expressed CD27 (another TNF family cell), demonstrated an increased risk of metastasis compared to CD27-cells, while the opposite relationship held for CD40-cells.

#### Nodal Metastasis

Several noteworthy interactions for nodal metastasis were noted and include: 1) CD34-PD-L1 interaction in the intratumoral region (CD34 positively associated with metastasis for PD-L1-cells and negatively associated for PD-L1+ cells), 2) an interaction between CD3 and CD44 at the tumor interface (high CD44 expression related to nodal metastasis for tumors with cytotoxicity (e.g., CD3+/CD8+) at the interface), and 3) the positive association between CD127 and nodal metastasis was strengthened for CD66b+ cells at the interface (**Figure 5**).

**Figure 5:**
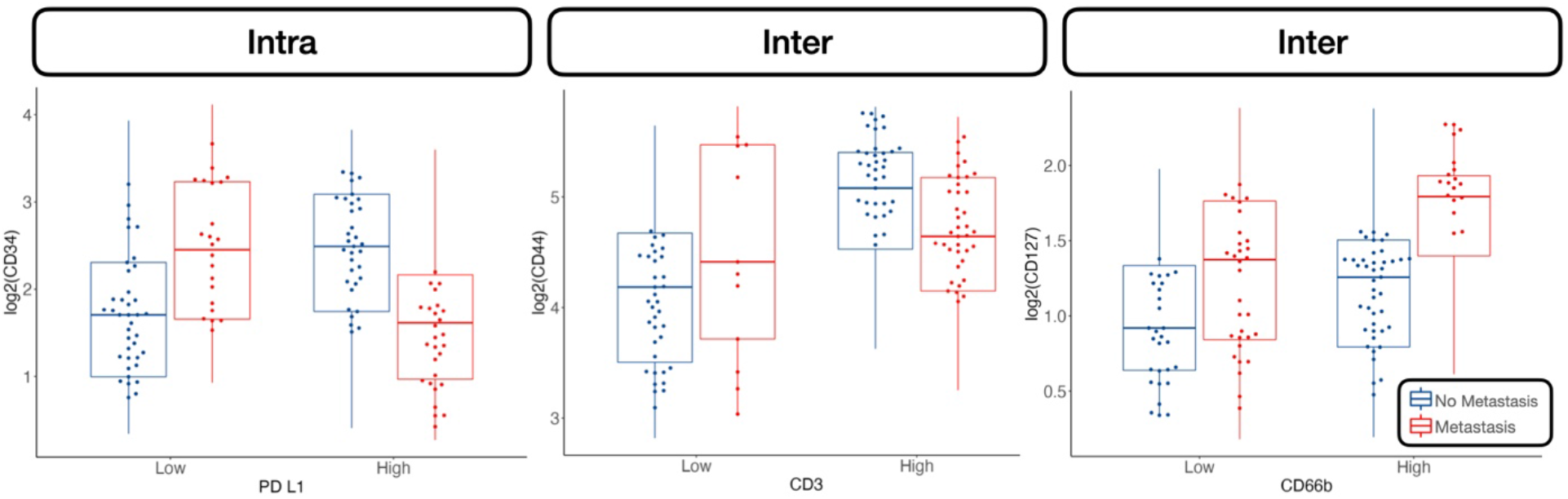
Select protein marker expression, conditional on cell type (stratified by median expression), predictive of nodal metastasis: A) CD34 expression stratified by PD-L1 expressing cells inside the tumor, B) CD44 stratified by CD3 at the *interface*, C) CD127 stratified by CD66b at the *interface*. Marker expression plotted in beeswarm plots were filtered based on the detection of outliers using a modified Tukey outlier test– after this initial filtering, only points between the 10% and 90% quantiles for each stratum were included

#### Distant Metastasis

For distant metastasis, increased PanCk expression for CD66b-cells was positively associated with distant metastasis. At the interface, increased expression of fibronectin was associated with metastasis, but only for CD66b+ cells. Away from the tumor, CD66b expression was correlated with metastasis for immune cells deficient in FAP-alpha expression (**Figure 6**).

**Figure 6:**
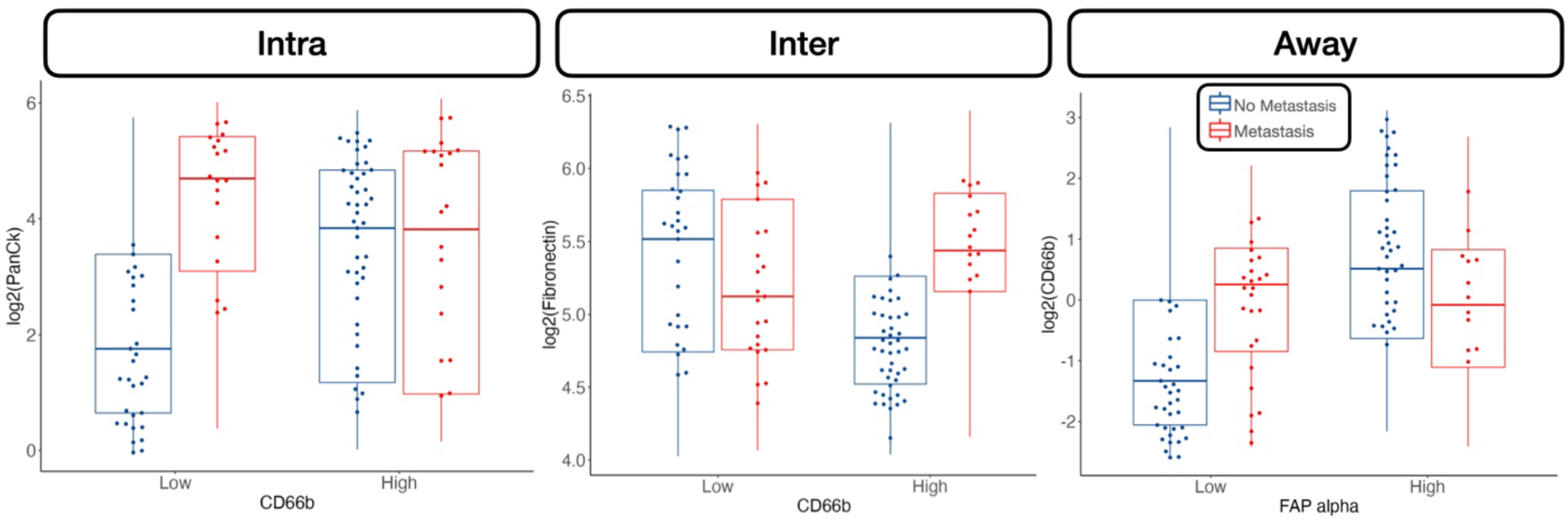
Select protein marker expression predictive of distant metastasis, conditional on cell type (stratified by median expression): A) PanCk expression stratified by CD66b expressing cells inside the tumor, B) Fibronectin stratified by CD66b at the *interface*, C) CD66b stratified by FAP-Alpha away from the tumor. Marker expression plotted in beeswarm plots were filtered based on the detection of outliers using a modified Tukey outlier test– after this initial filtering, only points between the 10% and 90% quantiles for each stratum were included

A complete listing of relevant metastasis-related interactions can be found in the Shiny app and supplementary table (**Supplementary Table 1**) and figures (**Supplementary Figures 7-10**).

### Differential Coexpression

#### Nodal metastasis

*Intratumoral:* An analysis of differential coexpression within architectures revealed conserved coexpression/colocalization amongst helper, cytotoxic T-cells and their co-activators (e.g., CD3, CD4, CD8, CD40) within the intratumoral region between patients with and without nodal metastasis. Important co-expressed genes did not change across tissue architectures. Fibronectin, CD44, FAP-alpha, and CD127 exhibited significant differential intratumoral coexpression between patients based on their lymph node status. *Interface/Away:* Interestingly, coexpression with fibronectin, FAP-alpha, and CD44 became increasingly less relevant (as defined by either differential or specific coexpression) for nodal metastasis at the interface and away from the tumor, while CD163 demonstrated the opposite relationship (**Figure 6; Supplementary Figures 11-15**).

#### Distant metastasis

Patterns of conserved coexpression are similar between patients with nodal and distant metastasis as compared to the controls. *Intratumoral:* However, coexpression with CD40 and ICOS became increasingly less relevant as a function of distance to the tumor for patients with distant metastasis, while CD27 coexpression became increasingly relevant with distance. Differential coexpression with fibronectin in the tumor’s periphery (*inter, away*) was more associated with distant metastasis than coexpression inside the tumor. *Interface/Away:* However, coexpression with fibronectin and FOXP3 at the periphery and beyond was not highly specific to metastasis/controls compared to intratumoral regions (**Figure 7; Supplementary Figures 16-18**).

**Figure 7:**
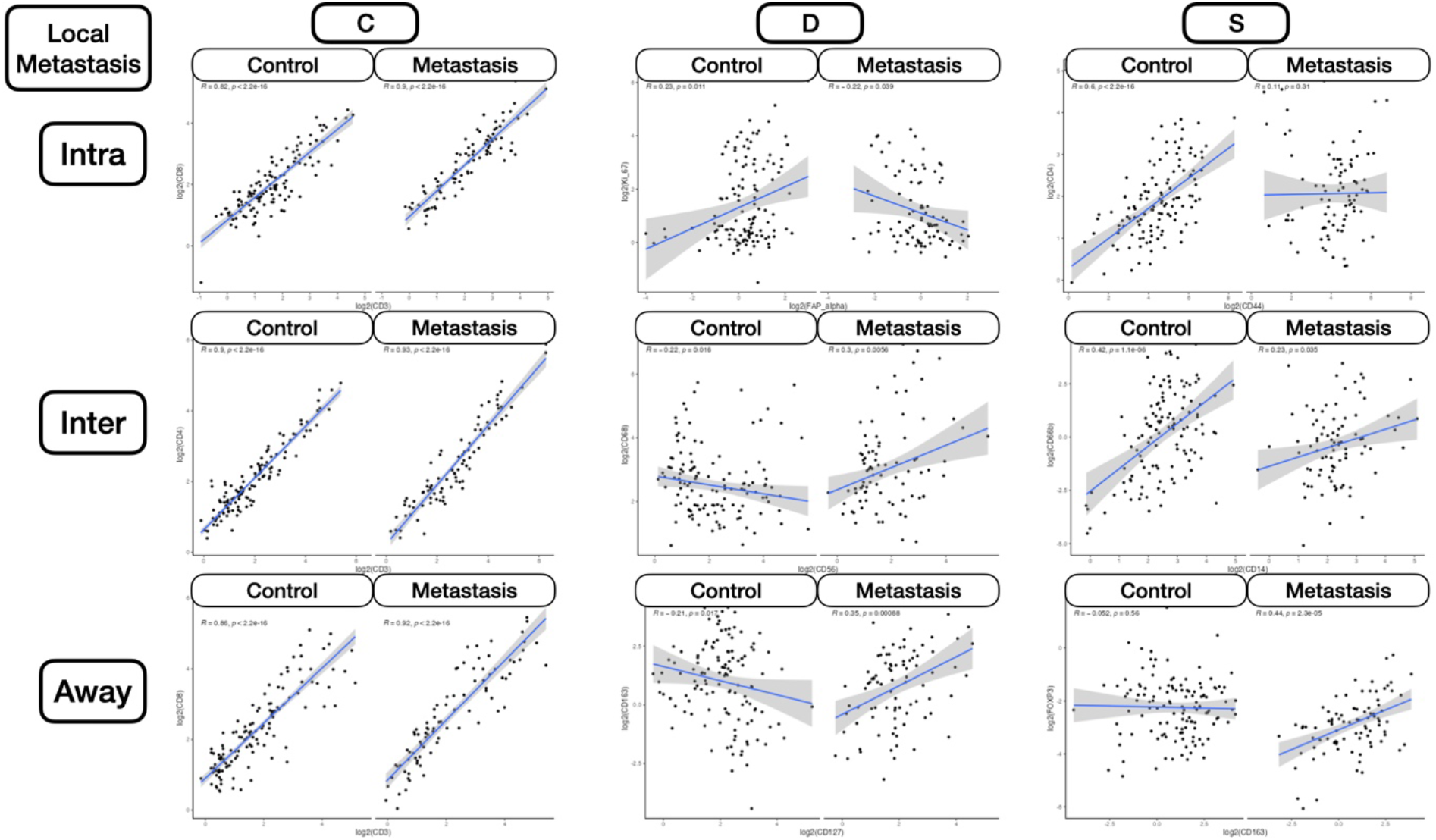
Differential Coexpression between select protein markers, stratified by lymph node metastasis status within three tissue architectures (*intra, inter, away*). C indicates whether coexpression was conserved between patients with and without metastasis; D indicates whether coexpression differed between patients with and without metastasis; S indicates whether significant coexpression was specific to either patients with or without metastasis

Quantitative findings can be found in the Shiny application. Significant markers were additionally predictive when taken together using hierarchical clustering (**Supplementary Figure 19**).

## Discussion

Examination of regional lymph nodes at the time of surgical resection is essential for CRC prognostication through accurate TNM staging. While it is important to maximize the number of lymph nodes assessed, recent population-based studies have shown that examination and processing of lymph node involvement is usually incomplete or inadequate. This incomplete or inadequate assessment can impact the accuracy of tumor staging and downstream disease management options, such as whether the patient should receive adjuvant chemotherapy. In response, it has been commonly suggested that methods which can better look for lymph nodes (e.g., submit higher percentage of resected fat) can increase lymph node yield. To complement these increasingly thorough assessments, developing alternative methods which assess lymph node involvement through indirect molecular mechanisms could be useful in cases where lymph node examination/processing is inadequate.

Multiplexed spatial transcriptomics and proteomics methods help identify novel predictive -omics signatures of metastasis through indirect observation from the primary site. In a set of stage pT3 tumors with or without nodal and/or distant metastases, we sought to identify spatial proteomic markers of metastasis with digital spatial profiling of immune cells. Furthermore, compared to previous studies with limited multiplexing capacity, we leveraged machine learning technologies to evaluate multiple markers in conjunction for increased capacity to determine metastasis status. We assessed three distinct architectures (*intra, inter, away*) within the primary site for 1) relative abundance (ratio tests), 2) interactions (classifier), and 3) coexpression (differential coexpression analysis). Our assessments revealed potential nodal and distant metastasis biomarkers separately and together for patients with and without tumor metastasis which we will pursue in the future using with additional data collection and orthogonal validation on assays less onerous and subject to variation as compared to the DSP.

While the limited sample of this study size precludes any firm conclusions, we demonstrated important trends of concordant expression levels across the distinct tumor architectures. Several proteins (e.g., GZMB, CD8, PD-L1, CD3, FOXP3, CD56, fibronectin, and CD66b) appeared to be important in metastasis. We identified several emergent trends that will motivate future work: 1) architectural differences, 2) the role of GZMB and the dual role of extracellular remodeling, 3) the role of PD-L1 expressing dendritic cells, 4) T-cell exhaustion in tumors with mismatch repair deficiencies, 5) paradoxical role of immune suppression in CRC, 6) predictive value of NK Cells in a non-immunosuppressive environment, and 7) neutrophil infiltrates and fibroblast activation.

### Architectural Differences

Our findings differed between the three distinct architectures. For instance, a CD34-PD-L1 interaction was observed in the intratumoral region. An interaction between CD3 and CD44 was found at the tumor interface. A positive association between CD127 and nodal metastasis was strengthened for CD66b+ cells at the interface. Fibronectin, CD44, FAP-alpha, and CD127 exhibited significant differential intratumoral coexpression between patients based on their lymph node status, whereas several helper, cytotoxic T-cells and their co-activators such as CD3, CD4, CD8, CD40 exhibited conserved coexpression within the intratumoral region between patients with and without nodal metastasis. These are examples of many architectural differences, reaffirming the importance of evaluating the tumor invasive margin/interface.

### Role of GZMB

GZMB is a potent molecule used by CD8 T cells to induce cytotoxicity when detecting certain antigens. As an extracellular matrix agent that induces apoptosis, GZMB is involved in cleaving certain target proteins, leading to DNA fragmentation and loss of membrane integrity ^43^. Its role in colorectal cancer was recently elucidated by Daemen et al. The authors found that GZMB is associated with a better prognosis of CRC both in MSI positive and negative clones. Its function was not only linked to cytotoxic T cells but was found to be elicited by tumor cells independent of CD8, with a role in cleaving vitronectin, fibronectin, and laminin ^44^. This hypothesis is supported by previous experiments of pre-treating the laminin matrix with GZMB with significant inhibition of cell spreading in the LIM1215 colon cancer cell line ^45^. We found that in dMMR patients, GZMB-expressing immune cells were associated with distant metastasis, which contradicts these prior findings. We also note in our study that fibronectin expression appeared to be positively associated with distant metastasis in immune-cells which did not express GZMB. While GZMB may assist in extracellular matrix (ECM) remodeling, allowing for the transmigration of T cells, similarly remodeling of the ECM could potentially facilitate the migration of tumor cells, suggesting a dual role for this protein ^46^.

### Role of PD-L1 expressing dendritic cells

PD-L1 is a protein found in the cell surface of tumor cells that couples with the PD-1 protein of T cells, causing inhibition of the T cells’ immune functions against the tumor. Therefore, our results, showing increased PD-L1 expression near immune cells at the invasive margin for local lymph node involvement and inside the tumor in dMMR patients for distant metastasis, agree with previous studies that link this molecule to immune evasion ^47^. Our study also agrees with a study of 221 patients with stage pT3 colon cancer that investigated cancer tissues immunostained to examine the prognostic impact of CD11c^+^ dendritic cell co-expressing PD-L1 and their spatial relationship with CD8^+^ T-cells. The authors found significant survival benefits for patients with intratumoral CD8^+^ cell density, stromal CD11c^+^ cell density, intratumoral CD11c^+^ PD-L1^+^ cell density, and stromal CD11c^+^ PD-L1^+^ cell density. CD8^+^ cell density was positively correlated with both CD11c^+^ cell density and CD11c^+^ PD-L1^+^ cell density in tumor epithelium and stromal compartments. CD11c is a member of the integrin family (adhesion molecule), and it is particularly expressed in dendritic cells ^48^. Dendritic cells are antigen-presenting cells that play an important role in adaptive immunity by attracting naïve T cells to activate, differentiate and finally infiltrate tumors. Studies show mature tumor-infiltrating dendritic cells to be associated with better prognosis ^49^. The value of CD11c dendritic cells in cancer control has been shown in various types of cancer. Wang et al. studied CD11c expression in gastric cancer and found that patients with high expression levels had, on average, a better survival and significantly reduced risk of relapse. Lee et al. studied tissue micro-arrays from 681 pre-treated patients with triple-negative breast cancer. Microscopically, CD11c cells are concentrated in areas with high numbers of TILs. Tumors with high expression of CD11c also had higher histological grades. More importantly, those with lymph node metastasis and high CD11c expression showed a trend of increased recurrence-free survival and significantly better overall survival (p=0.047) compared to those with low CD11c expression^50^. These results are also corroborated by findings in our paper, where PD-L1 expressing CD11c+ cells (dendritic cells) were negatively associated with distant metastasis.

### T-cell exhaustion in tumors with mismatch repair deficiencies

Stimulating the recruitment of cytotoxic T cells has been clinically explored in immunotherapy. One example of a molecule that has been studied as a potential therapeutic target is CD3, a protein found on the surface of T cells, with downstream signaling resulting in the activation of T-cells. Evidence suggests CD3 is an important target as it can redirect T cells to attack tumors by secreting inflammatory cytokines and cytolytic molecules, but overproduction may cause a cytokine storm, associated with worse outcomes. CD3 is an important pan-T-cell antigen– direct targeting of CD3 cells can redirect other T cells to attack the tumors through secreting inflammatory cytokines and cytolytic molecules ^50–52^. We noticed that expression of CD3 and CD8 inside the tumor and at the invasive margin were positively associated with local metastasis, which contradicts the general notion that the presence of these biomarkers suggests a favorable diagnosis. This is not the first-time contradictions of these predictive effects have been discussed, as it has been reported that these markers’ predictive value can vary widely based on mismatch repair status ^53^. This is partly because a mismatch repair deficient tumor could exhibit a T-cell exhaustion phenotype, even prior to metastasis, which could indicate a diminished capacity to impede metastasis (e.g., exhaustion, functional suppression from other TME immune cells). In particular, the association with CD8 was statistically significant for pMMR patients and marginally significant for dMMR patients. As tumors were not profiled prior to metastasis, reverse causality is also possible (i.e., infiltration at the primary site as instigated by metastasis). As the Immunoscore, a digital pathology assay which assesses immune cells at the tumor’s core and invasive margin, is derived from these CD3 and CD8 measurements, further exploration of the interaction between the Immunoscore and mismatch repair for colon cancer prognostication is warranted ^7,54^. Patients with dMMR also exhibit heterogenous immune activation (e.g., COLD and HOT tumors), which are related to different prognostic outcomes and can be elucidated through whole transcriptomic characterization ^55^.

### Paradoxical role of immune suppression in CRC

The FOXP3 gene, located on the X chromosome, is a transcription factor that plays an important role in the production of CD4+ CD25+ regulatory T cells (Tregs) ^56^. However, its exact role in cancer development and metastasis remains elusive. While in the past FOXP3 expression had been associated only with regulatory T cells and studied almost exclusively in lymphoid tissue, FOXP3 expression in nearby neighboring tumor cells can lead to immune suppression by curbing the immune response, further complicating the conclusion of previous research results ^57^. While for most human carcinomas, FOXP3 is typically associated with poor prognosis, paradoxically, the opposite holds true for CRC ^58,59^. FOXP3 can also function as a tumor suppressor, with its expression linked to better outcomes ^60^. However, the opposite has also been reported in other cancers, namely its action as an oncogene through activation of the APC and epithelial to mesenchymal pathways through fibroblast differentiation (release of GZMB) ^61^. Further complicating the analysis of FOXP3 is the fact that it has four isoforms, each being functional but interacting with different molecules that play a role in cancer progression. For instance, further evidence suggests that FOXP3-expressing T-regulatory cells could be further fractionated into lineages, which are non-suppressive but secrete pro-inflammatory cytokines ^62^. Several studies also suggest that the protective effects of FOXP3 may be mediated through, interact with, or be impacted by the abundance of certain host microbiota by suppressing the inflammatory response to gut microbiota. Our study noted the protective effects of FOXP3 away from the tumor, particularly for dMMR patients. Regardless, more studies are needed, controlling for FOXP3’s isoforms and their various interactions in the tumor and its microenvironment ^63^.

### Prognostic value of NK Cells in a non-immunosuppressive environment

Natural Killer (NK) cells infiltrate solid tumors to lyse cancerous cells. Our study identified CD56+ NK cells as a significant protective factor against metastasis in the peritumoral region, but only for dMMR patients. This is consistent with the observation of substantial inhibition of NK in an immunosuppressive TME, where presence, despite the inhibition, would prove favorable. Sconochia et al. studied NK cells in 1410 CRC specimens and other solid tumors and mostly confirmed that NK cells are minimally detectable. However, the authors identified an interesting subgroup of patients that exhibited a high degree of NK cell infiltration along with their CD8+ T cells. CD56 was used as the antigenic biomarker of NK cells. Using CD56 expression to track the degree of NK cell infiltration in colorectal tumors and further characterizing CD8+ lymphocytes, they found that CD56+ CD8+ patients presented significantly higher survival (80%) vs. 55% for the CD56-CD8+ group ^64^. Similar results were shown by Alderdice et al., with CD56+ rectal cancer patients experiencing significantly better overall survival. The authors propose CD56+ as a signature for clinical decision-making and treatment duration. Others have explored how NK cells may be activated after treatment with cetuximab plus IL-2 or IL-15 ^65^. We have identified a potential interaction between CD56 and MMR status.

### Neutrophil infiltrates and fibroblast activation

CD66b is an antigenic marker of tumor-infiltrating neutrophils, representing proinflammatory myeloid infiltrates ^66,67^. Infiltrating neutrophils seem to increase cancer invasion, lymph node metastasis, and tumor stage ^68,69^. The neutrophil-to-lymphocyte ratio (NLR) in the intratumoral regions commonly serves as a prognostic indicator. Prior studies have suggested that a high NLR ratio could suggest a poor prognosis. In CRC, the prognostic role of tumor-associated neutrophils has not been established since various authors report conflicting results ranging from poor prognosis, to no association, to improved prognosis. For instance, similar to our study findings, a prior study suggested that absence of CD66b+ immune cell infiltrates could be predictive of local metastasis ^70^. It has been implied that favorable antitumoral effects could be mediated through costimulation of other cell lineages, e.g., CD3+/CD8+. There is evidence to suggest cross-talk between cytotoxic T cells and neutrophils could be further substantiated by studying factors pertaining to neutrophil recruitment. Interestingly, more recent publications in a comprehensive literature review tend to report neutrophils as a protective factor, whereas older articles demonstrate a negative role ^71,72^. We also noted the potential interaction between CD66b+ cells, fibronectin, and other fibroblast activation proteins (e.g., FAP-Alpha). Fibronectin has been previously implicated as means to promote invasion and metastasis and is a crucial structural component of angiogenic tumors. At the invasive margin, we noticed that fibronectin’s impact on local metastasis was specific to CD66+ cells as compared to CD66-cells. These effects were not identified away from the tumor. However, the opposite effect was noted for FAP-alpha– non-fibroblast activating (FAP-Alpha-) CD66+ cells away from the tumor exhibited a higher risk of metastasis as compared to FAP-alpha-CD66-cells ^73^. For CD66b+ cells in the *away region* (e.g., stroma), FAP-alpha appeared to be negatively associated with local involvement. We also noticed colocalization between these two cell types away from the tumor ^74^. These findings coincide with prior research suggesting that cancer-associated fibroblasts can regulate the recruitment of myeloid cells ^75,76^.

There are several study limitations worth mentioning. The sample size was limited due to the high cost of spatial proteomics assays on a per-slide basis, making it challenging to control for tumor site. The sample size also limits both the certainty of our study findings as well as the power to reveal additional biomarkers of metastasis. As another example, we did not assess tumors with distant but not local metastasis, which may derive separate predictive cell lineages as the notion of lymph node metastasis serving as a “staging ground” for distant metastasis has been disputed as the exclusive progression ^77–80^. We attempted to control for potential batch effects by balancing our batches with an even number of patients with and without metastasis and further adjustments via mixed effects machine learning and statistical models. Even so, there may be technical factors introducing heterogeneous expression, which were uncontrolled. Prior treatment (e.g., chemotherapy) and comorbidities may have interfered with the assessment of the primary though this is something we exclusively searched and filtered for as inclusion criteria. The nCounter proteomics assay does not account for different protein isoforms and post-translational modifications (e.g., phosphorylation), which could prove additionally predictive. Furthermore, while many of the ROIs placed away from the tumor were placed in the stroma, we did not explicitly account for additional tissue architectural differences (e.g., peritumoral fat, immune nests, benign epithelium, etc.). While we had developed a semi-autonomous workflow for placement of ROIs, biased placement of DSP ROIs may impact study findings, warranting the exploration of assays which do not suffer from these effects (e.g., Visium Spatial Transcriptomics) ^81^. The Visium assay could also provide the capacity for untargeted characterization of the whole transcriptome at 50-micron resolution. Pairing these assays with a proteomics assessment will be explored in future work. Promising biomarkers established from these studies will be orthogonally validated through immunostaining.

Despite these limitations, leveraging the Digital Spatial Profiler for a highly multiplexed assessment of colorectal tumors has allowed for the simultaneous exploration of several emergent pathways and immunomodulatory effects which characterize the potential for tumor metastasis. Consulting the known literature on TIME to corroborate our study conclusions has demonstrated that CRC tumor immunology is still riddled with contradictory findings that continue to perplex researchers on this important subject. Spatial profiling provides an opportunity to further disaggregate the TIME. With the advent of multimodal spatial assays, which allow for greater multiplexing at higher resolution, future iterations of these studies may further elucidate the precise mechanisms of metastasis as means to develop low-cost diagnostic/prognostic tests that complement traditional assessments (e.g., Immunoscore, lymph node assessment) and inform novel therapeutics.

## Conclusion

Deciphering the Tumor Immune Microenvironment is key to improving the prognostication and treatment of colorectal adenocarcinoma. Spatial assessment technologies will continue to play a key role in delineating crucial architectural and cellular components of CRC tumor metastasis. We utilized the Digital Spatial Profiler to uncover spatial proteomics biomarkers of nodal and distant metastasis. Our study identified proteomics markers that can provide additional predictive value for metastasis when used in conjunction (e.g., relative abundance, coexpression, interaction, etc.). There is significant room for further investigation through unbiased and untargeted spatial RNA assays, as emergent themes appear contradictory to components of the previous literature. In the future, we plan to leverage high dimensional spatial mRNA and single-cell assays to further contextualize metastasis etiology and pathogenesis and to operationalize significant results into informative tests which complement existing assessment methods for CRC prognosis.

## Data Availability

Study data can be explored using an Rshiny application, available at the following URL: https://levylab.shinyapps.io/ViewColonDSPResults (user: edit_user, password: cgat2022). Complete set of data produced in the present study are available upon reasonable request to the authors.

## Acknowledgements

We would like to thank Gabriel Brooks and Linda Vahdat for their thoughtful discussions of the subject matter. This work was supported by NIH grants R01CA216265, R01CA253976, and P20GM104416 to BC, Dartmouth College Neukom Institute for Computational Science CompX awards to BC, JL and LV, and DCC, DPLM Clinical Genomics and Advanced Technologies EDIT program. JL is supported through NIH subawards P20GM104416 and P20GM130454. The funding bodies above did not have any role in the study design, data collection, analysis and interpretation, or writing of the manuscript.

## Notes

**Funding Support:** NIH subawards P20GM104416 and P20GM130454

### Competing Interest Statement

The authors have declared no competing interest.

### Author Declarations

IRB of Dartmouth Health gave ethical approval for this work.

