## Supplementary Materials for "Identification of Spatial Proteomic Signatures of Colon Tumor Metastasis: A Digital Spatial Profiling Approach"

### Differential Expression Supplementary Figures and Tables

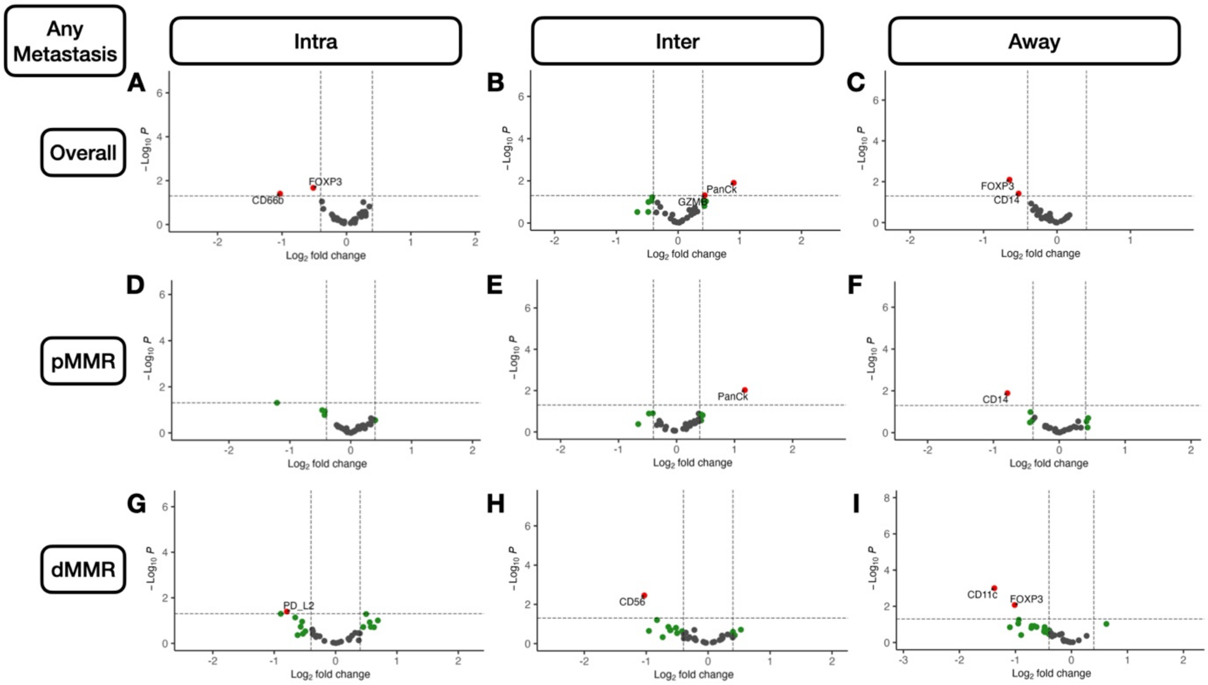


**Supplementary Figure 1: Volcano Plots Illustrating Differentially Expressed Protein Markers of Any Metastasis:** A-I) Results stratified by tissue architecture: A,D,G) *Intratumoral*; B,E,H) *peritumoral*; C,F,I) *away/stroma*; results also stratified by MMR-status: D-F) *pMMR*; G-I) *dMMR*; statistical significance cutoff at $\alpha = 0.05$; x-axis indicates effect size and directionality (positive x-value indicates metastasis-related marker; negative indicates decreased metastasis risk); y-axis indicates effect significance (positive y-value indicates lower p-value)

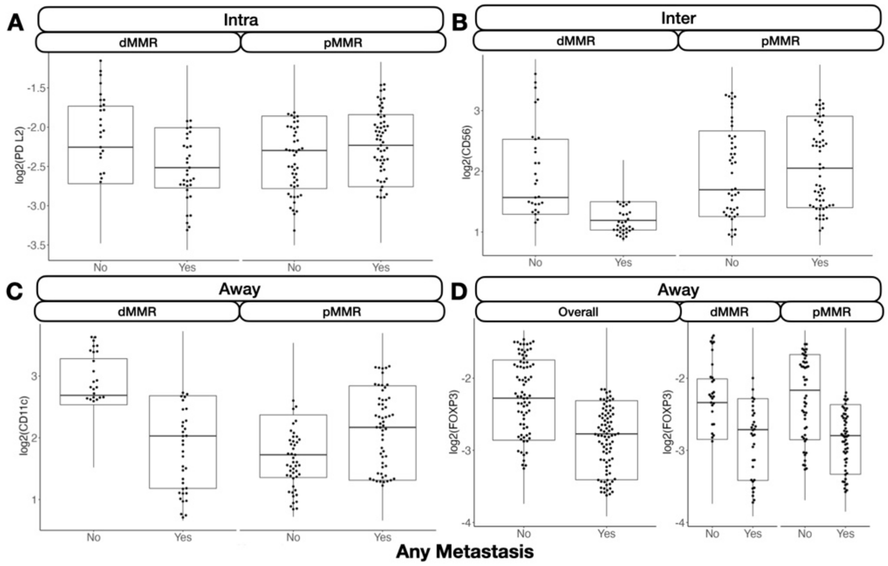


**Supplementary Figure 2: Boxplots and beeswarm scatterplots of select protein marker expression for biomarkers predictive of any metastasis, stratified by MMR-status:** A) PD-L2 inside the tumor; B) CD56 at the *interface*; C) CD11c away from tumor; D) FOXP3 away from tumor. Marker expression plotted in beeswarm plots were filtered based on detection of outliers using a modified Tukey outlier test– after this initial filtering, only points between the 10% and 90% quantiles for each stratum were included

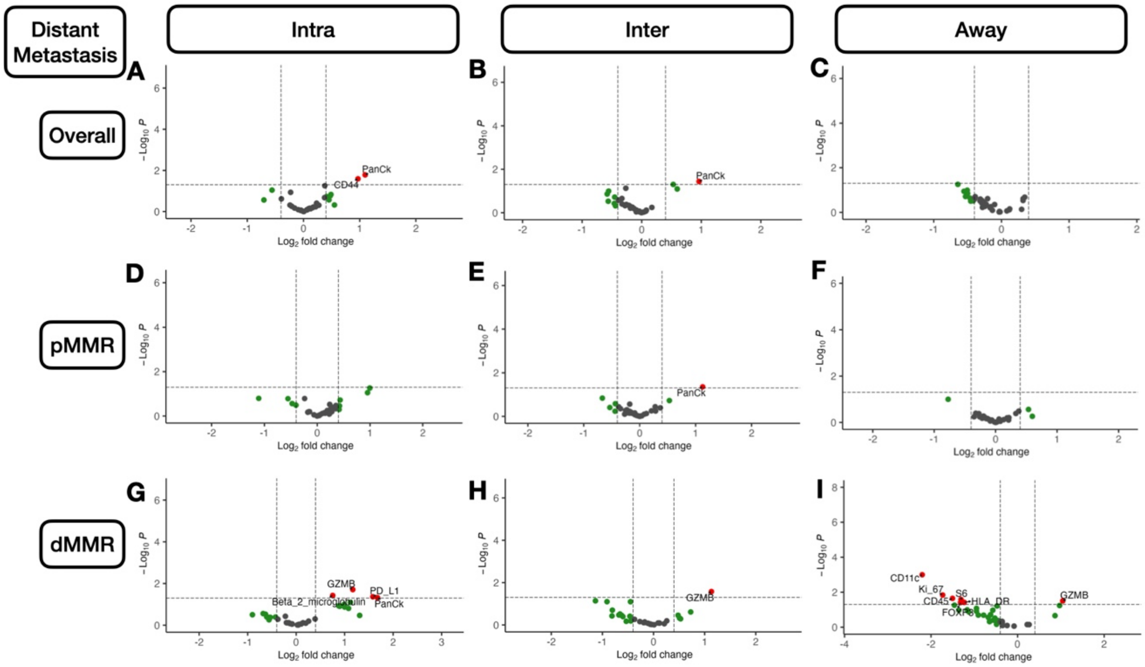


**Supplementary Figure 3:** **Volcano Plots Illustrating Differentially Expressed Protein Markers of Distant Metastasis:** A-I) Results stratified by tissue architecture: A,D,G) *Intratumoral*; B,E,H) *peritumoral*; C,F,I) *away/stroma*; results also stratified by MMR-status: D-F) *pMMR*; G-I) *dMMR*; statistical significance cutoff at $\alpha = 0.05$; x-axis indicates effect size and directionality (positive x-value indicates metastasis-related marker; negative indicates decreased metastasis risk); y-axis indicates effect significance (positive y-value indicates lower p-value)

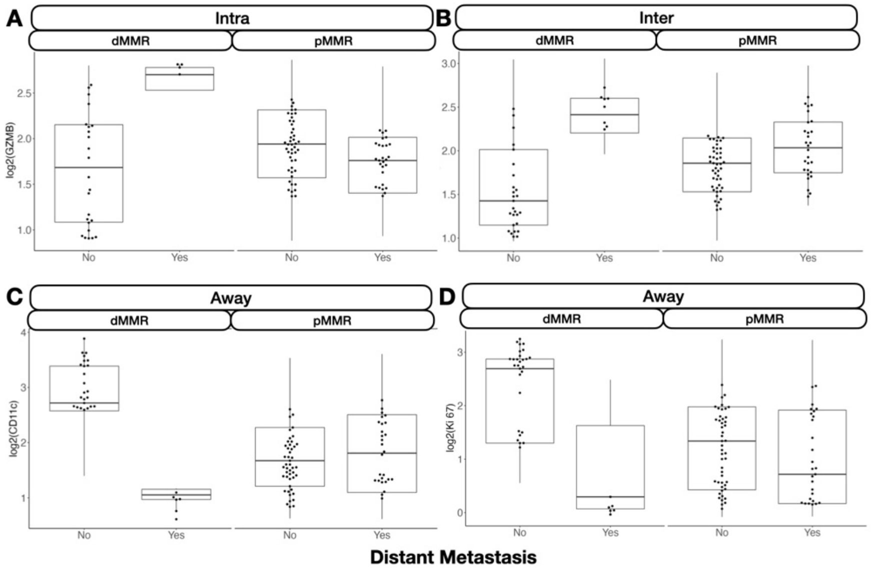


**Supplementary Figure 4:** **Boxplots and beeswarm scatterplots of select protein marker expression for biomarkers predictive of distant metastasis, stratified by MMR-status:** A) GZMB inside the tumor;

B) GZMB at the *interface*; C) CD11c away from tumor; D) Ki-67 away from tumor. Marker expression plotted in beeswarm plots were filtered based on detection of outliers using a modified Tukey outlier test– after this initial filtering, only points between the 10% and 90% quantiles for each stratum were included

### Differential Ratio Supplementary Figures and Tables

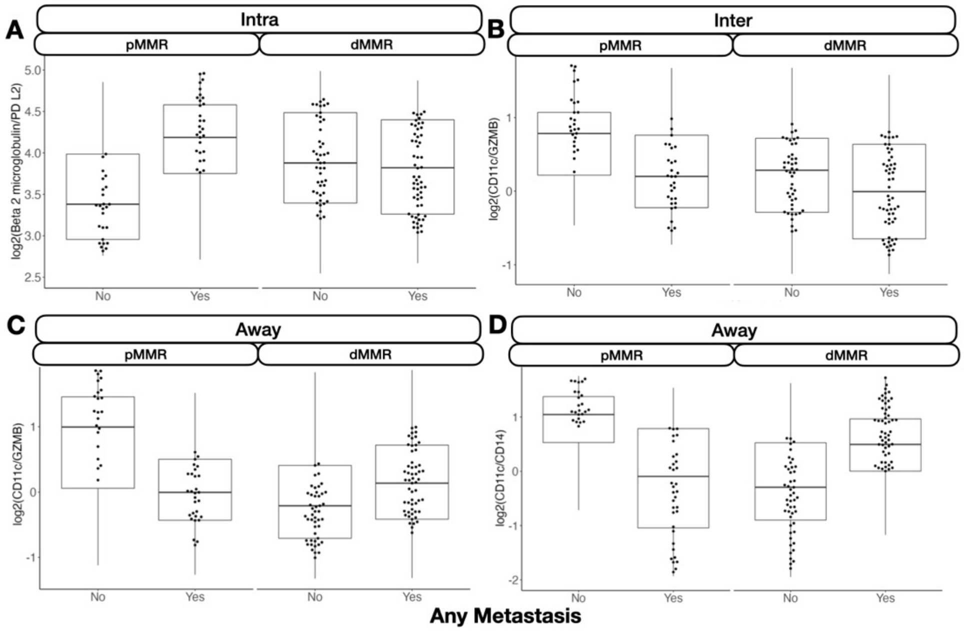


**Supplementary Figure 5: Boxplots and beeswarm scatterplots of relative protein expression between two markers, predictive of any metastasis, stratified by MMR-status:** A) Beta-2-microglobulin/PD-L2 inside the tumor; B) CD11c/GZMB at the *interface*; C) CD11c/GZMB away from tumor; D) CD11c/CD14 away from tumor. Marker expression plotted in beeswarm plots were filtered based on detection of outliers using a modified Tukey outlier test– after this initial filtering, only points between the 10% and 90% quantiles for each stratum were included

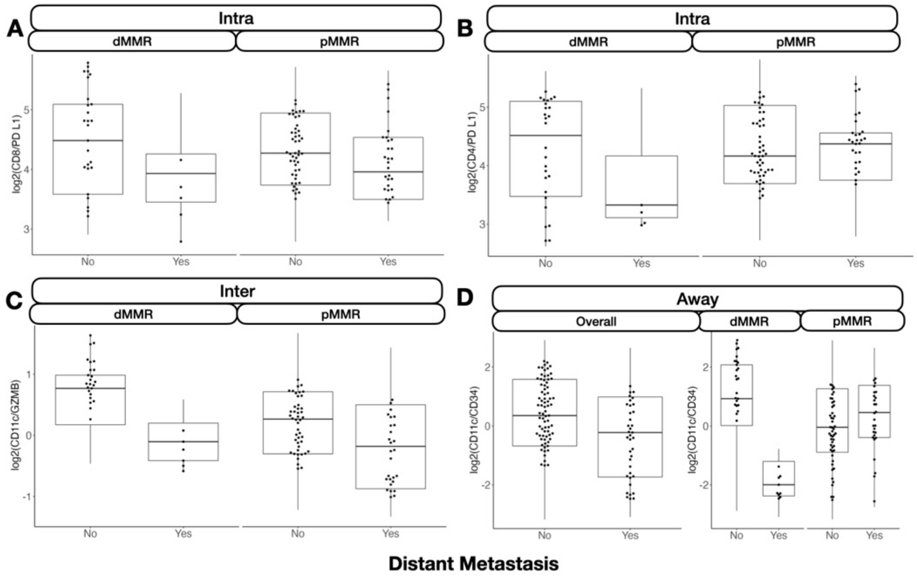


**Supplementary Figure 6: Boxplots and beeswarm scatterplots of relative protein expression between two markers, predictive of distant metastasis, stratified by MMR-status:** A) CD8/PD-L1 inside the tumor; B) CD4/PD-L1 inside the tumor; C) CD11c/GZMB at the interface; D) CD11c/CD34 away from tumor. Marker expression plotted in beeswarm plots were filtered based on detection of outliers using a modified Tukey outlier test– after this initial filtering, only points between the 10% and 90% quantiles for each stratum were included

### Interaction Supplementary Figures and Tables

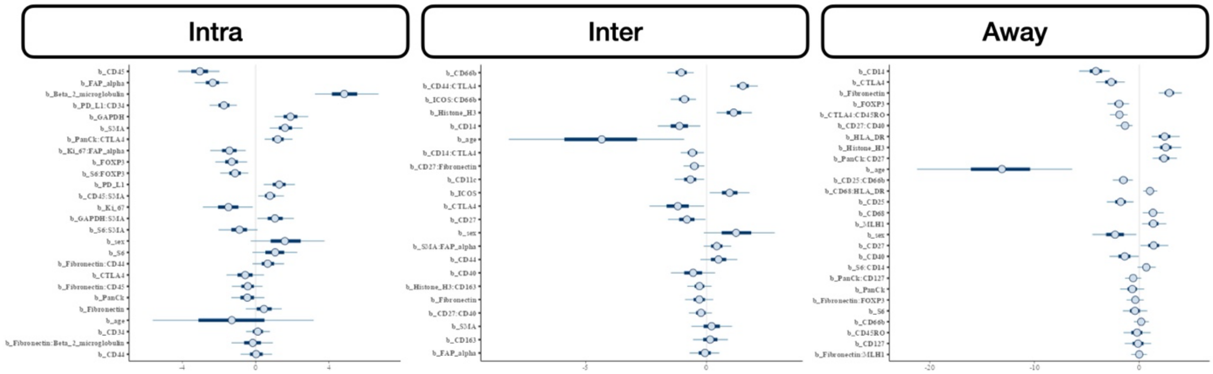


**Supplementary Figure 7: Posterior interval estimates from MCMC draws from Bayesian generalized linear mixed effects models for prediction of any metastasis** from standardized protein markers and their interactions, stratified by architecture; predictors were derived from MEML models and subselected using Horseshoe LASSO prior to unpenalized statistical testing

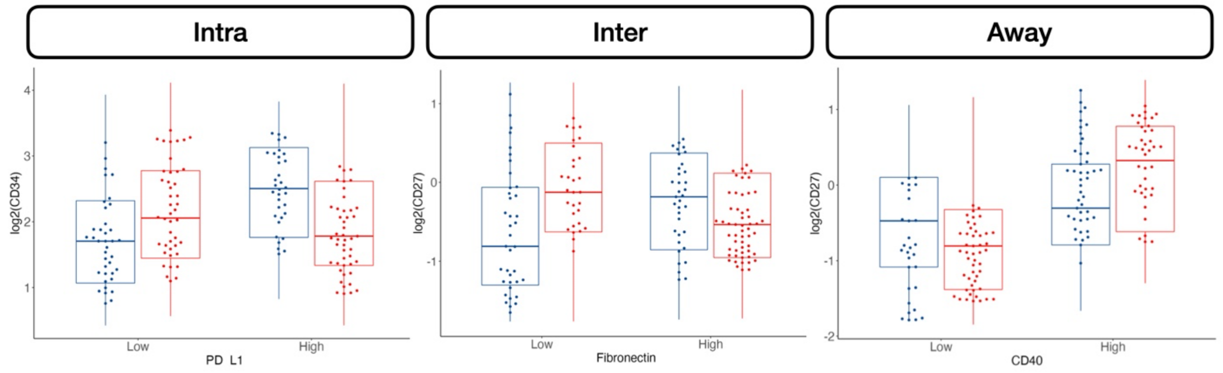


**Supplementary Figure 8: Boxplots and beeswarm scatterplots of select protein marker expression, conditional on cell type (stratified by median expression), predictive of any metastasis:** A) CD34 expression stratified by PD-L1 expressing cells inside the tumor, B) CD27 stratified by Fibronectin at the *interface*, C) CD27 stratified by CD40 at the *interface*. Marker expression plotted in beeswarm plots were filtered based on detection of outliers using a modified Tukey outlier test– after this initial filtering, only points between the 10% and 90% quantiles for each stratum were included

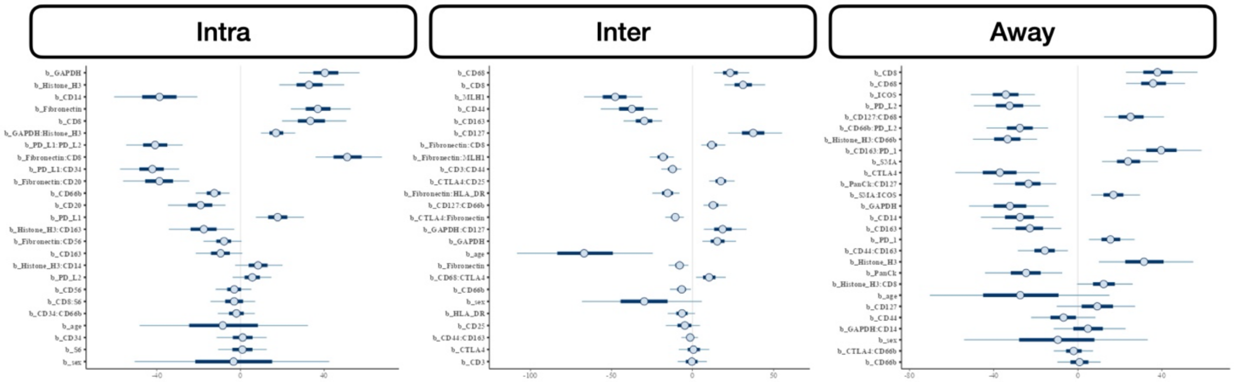


**Supplementary Figure 9: Posterior interval estimates from MCMC draws from Bayesian generalized linear mixed effects models for prediction of nodal metastasis** from standardized protein markers and their interactions, stratified by architecture; predictors were derived from MEML models and subselected using Horseshoe LASSO prior to unpenalized statistical testing

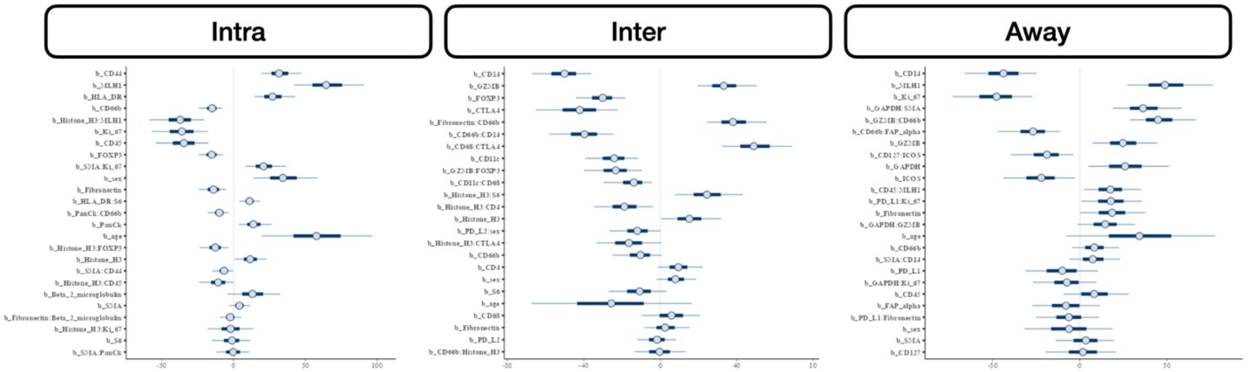


**Supplementary Figure 10: Posterior interval estimates from MCMC draws from Bayesian generalized linear mixed effects models for prediction of distant metastasis** from standardized

protein markers and their interactions, stratified by architecture; predictors were derived from MEML models and subselected using Horseshoe LASSO prior to unpenalized statistical testing

**Supplementary Table 1: Posterior 95% credible interval estimates and p-values from MCMC draws from Bayesian generalized linear mixed effects models for prediction of any, nodal, and distant metastasis** from standardized protein markers and their interactions, stratified by architecture; predictors were derived from MEML models and subselected using Horseshoe LASSO prior to unpenalized statistical testing

| Target | Macroarchitecture | Marker | log(OR) | 2.5% CI | 97.5% CI | p |
|---|---|---|---|---|---|---|
| **Any Metastasis** | Intra | CD45 | -3.039 | -4.387 | -1.727 | <0.001 |
| | | FAP-alpha | -2.338 | -3.492 | -1.376 | <0.001 |
| | | Beta-2-microglobulin | 4.834 | 2.888 | 6.97 | <0.001 |
| | | GAPDH | 1.904 | 0.831 | 3.034 | <0.001 |
| | | PD-L1:CD34 | -1.721 | -2.606 | -0.849 | <0.001 |
| | | SMA | 1.609 | 0.531 | 2.692 | 0.001 |
| | | PanCk:CTLA4 | 1.214 | 0.344 | 2.132 | 0.003 |
| | | Ki-67:FAP-alpha | -1.422 | -2.663 | -0.392 | 0.007 |
| | | FOXP3 | -1.3 | -2.263 | -0.219 | 0.008 |
| | | S6:FOXP3 | -1.118 | -2.066 | -0.245 | 0.01 |
| | | PD-L1 | 1.292 | 0.303 | 2.354 | 0.014 |
| | | CD45:SMA | 0.785 | 0.007 | 1.663 | 0.04 |
| | | Ki-67 | -1.477 | -3.027 | 0.107 | 0.065 |
| | | GAPDH:SMA | 1.055 | -0.13 | 2.244 | 0.068 |
| | Inter | CD66b | -1.046 | -1.733 | -0.43 | <0.001 |
| | | CD44:CTLA4 | 1.501 | 0.832 | 2.181 | <0.001 |
| | | ICOS:CD66b | -0.915 | -1.581 | -0.329 | 0.002 |
| | | Histone-H3 | 1.125 | 0.302 | 2.039 | 0.01 |
| | | CD14 | -1.117 | -2.163 | -0.12 | 0.028 |
| | | age | -4.334 | -8.687 | -0.083 | 0.034 |
| | | CD14:CTLA4 | -0.583 | -1.153 | -0.008 | 0.04 |
| | | CD27:Fibronectin | -0.502 | -1.039 | -0.027 | 0.048 |
| | | CD11c | -0.668 | -1.398 | 0.054 | 0.059 |
| | | ICOS | 0.954 | 0.046 | 2.013 | 0.062 |
| | Away | CD14 | -4.141 | -5.986 | -2.517 | <0.001 |
| | | CTLA4 | -2.669 | -4.46 | -1.17 | <0.001 |
| | | HLA-DR | 2.416 | 0.839 | 4.023 | <0.001 |
| | | Fibronectin | 2.869 | 1.643 | 4.242 | <0.001 |
| | | FOXP3 | -1.918 | -3.273 | -0.806 | <0.001 |
| | | Histone-H3 | 2.526 | 0.997 | 4.161 | <0.001 |

| | | | | | | |
|---|---|---|---|---|---|---|
| | | CTLA4:CD45RO | -1.884 | -2.864 | -0.873 | <0.001 |
| | | PanCk:CD27 | 2.357 | 1.059 | 3.8 | <0.001 |
| | | CD27:CD40 | -1.355 | -2.331 | -0.485 | <0.001 |
| | | age | -13.115 | -21.787 | -4.843 | 0.001 |
| | | CD25:CD66b | -1.519 | -2.662 | -0.376 | 0.007 |
| | | CD68:HLA-DR | 1.023 | 0.232 | 1.855 | 0.01 |
| | | CD25 | -1.761 | -3.317 | -0.308 | 0.012 |
| | | CD68 | 1.312 | 0.132 | 2.551 | 0.026 |
| | | MLH1 | 1.342 | -0.003 | 2.769 | 0.037 |
| | | sex | -2.306 | -4.701 | 0.273 | 0.062 |
| **Lymph Node** | Intra | GAPDH | 40.372 | 26.076 | 59.699 | <0.001 |
| | | Histone-H3 | 32.753 | 14.779 | 51.232 | <0.001 |
| | | CD14 | -38.749 | -63.015 | -16.251 | <0.001 |
| | | Fibronectin | 36.971 | 22.062 | 55.224 | <0.001 |
| | | CD8 | 33.386 | 16.564 | 52.729 | <0.001 |
| | | GAPDH:Histone-H3 | 16.941 | 8.287 | 27.285 | <0.001 |
| | | PD-L1:PD-L2 | -40.812 | -56.708 | -24.94 | <0.001 |
| | | Fibronectin:CD8 | 51.059 | 32.692 | 69.931 | <0.001 |
| | | PD-L1:CD34 | -42.143 | -59.913 | -26.097 | <0.001 |
| | | Fibronectin:CD20 | -38.785 | -58.59 | -21.546 | <0.001 |
| | | CD66b | -12.529 | -22.818 | -3.713 | 0.001 |
| | | CD20 | -19.074 | -36.174 | -4.496 | 0.001 |
| | | PD-L1 | 17.74 | 5.273 | 32.625 | 0.005 |
| | | Histone-H3:CD163 | -17.559 | -37.257 | -0.27 | 0.048 |
| | Inter | CD68 | 23.221 | 10.674 | 36.463 | <0.001 |
| | | CD8 | 31.126 | 16.869 | 46.78 | <0.001 |
| | | MLH1 | -47.706 | -68.787 | -26.666 | <0.001 |
| | | CD44 | -37.423 | -59.379 | -18.241 | <0.001 |
| | | CD163 | -29.714 | -44.245 | -16.291 | <0.001 |
| | | CD127 | 37.405 | 18.879 | 57.735 | <0.001 |
| | | Fibronectin:CD8 | 11.835 | 3.626 | 20.96 | <0.001 |
| | | CTLA4:Fibronectin | -10.663 | -17.799 | -4.488 | <0.001 |
| | | Fibronectin:MLH1 | -18.175 | -27.639 | -10.125 | <0.001 |
| | | CD3:CD44 | -12.243 | -20.168 | -5.263 | <0.001 |
| | | CTLA4:CD25 | 17.457 | 8.946 | 27.191 | <0.001 |
| | | Fibronectin:HLA-DR | -15.313 | -25.618 | -6.054 | <0.001 |
| | | CD127:CD66b | 12.681 | 5.39 | 22.081 | <0.001 |
| | | GAPDH:CD127 | 18.595 | 4.351 | 34.623 | 0.004 |
| | | GAPDH | 15.321 | 4.077 | 28.716 | 0.006 |

| | | | | | | |
|---|---|---|---|---|---|---|
| | | age | -66.911 | -115.304 | -17.176 | 0.009 |
| | | Fibronectin | -7.959 | -15.123 | -0.717 | 0.01 |
| | | CD68:CTLA4 | 10.199 | -0.053 | 21.788 | 0.034 |
| | | CD66b | -6.664 | -15.117 | 0.012 | 0.046 |
| | Away | CD8 | 37.891 | 19.509 | 59.605 | <0.001 |
| | | CD68 | 35.811 | 20.207 | 53.675 | <0.001 |
| | | SMA | 23.877 | 9.421 | 41.247 | <0.001 |
| | | ICOS | -34.187 | -52.496 | -16.808 | <0.001 |
| | | CTLA4 | -36.971 | -61.637 | -14.037 | <0.001 |
| | | PD-L2 | -32.378 | -52.13 | -15.376 | <0.001 |
| | | PanCk:CD127 | -23.409 | -42.446 | -6.946 | <0.001 |
| | | CD127:CD68 | 25.061 | 10.195 | 43.892 | <0.001 |
| | | CD66b:PD-L2 | -27.541 | -44.953 | -10.444 | <0.001 |
| | | Histone-H3:CD66b | -33.359 | -51.725 | -16.151 | <0.001 |
| | | CD163:PD-1 | 39.624 | 20.767 | 61.984 | <0.001 |
| | | SMA:ICOS | 16.936 | 4.413 | 31.512 | 0.003 |
| | | GAPDH | -32.225 | -56.075 | -10.077 | 0.004 |
| | | CD14 | -27.47 | -48.672 | -7.584 | 0.005 |
| | | CD163 | -22.797 | -42.808 | -4.581 | 0.006 |
| | | PD-1 | 15.506 | 3.056 | 28.661 | 0.008 |
| | | CD44:CD163 | -15.615 | -29.423 | -1.475 | 0.01 |
| | | Histone-H3 | 31.433 | 4.702 | 57.547 | 0.015 |
| | | PanCk | -24.571 | -46.611 | -3.281 | 0.016 |
| **Distant** | Intra | CD44 | 31.691 | 16.236 | 49.245 | <0.001 |
| | | MLH1 | 64.486 | 37.258 | 94.808 | <0.001 |
| | | HLA-DR | 27.134 | 10.992 | 44.573 | <0.001 |
| | | Ki-67 | -35.992 | -59.266 | -12.567 | <0.001 |
| | | CD66b | -15.009 | -25.065 | -6.138 | <0.001 |
| | | Histone-H3:MLH1 | -37.184 | -60.805 | -16.201 | <0.001 |
| | | CD45 | -34.587 | -58.293 | -14.867 | 0.001 |
| | | FOXP3 | -15.282 | -26.159 | -6.366 | 0.002 |
| | | SMA:Ki-67 | 20.839 | 6.056 | 40.037 | 0.003 |
| | | sex | 34.28 | 11.175 | 63.672 | 0.005 |
| | | Fibronectin | -14.008 | -25.631 | -3.105 | 0.006 |
| | | HLA-DR:S6 | 11.09 | 2.464 | 19.418 | 0.008 |
| | | PanCk:CD66b | -9.889 | -19.021 | -1.47 | 0.014 |
| | | PanCk | 13.782 | 1.195 | 27.816 | 0.015 |
| | | age | 57.78 | 14.312 | 106.856 | 0.016 |
| | | Histone-H3:FOXP3 | -12.73 | -26.361 | -1.866 | 0.028 |

| | | | | | | |
|---|---|---|---|---|---|---|
| | Inter | CD14 | -50.12 | -69.995 | -33.198 | <0.001 |
| | | GZMB | 33.202 | 17.04 | 52.875 | <0.001 |
| | | FOXP3 | -30.155 | -45.085 | -15.273 | <0.001 |
| | | CD11c | -24.114 | -41.736 | -9.199 | <0.001 |
| | | CTLA4 | -42.173 | -67.718 | -17.716 | <0.001 |
| | | Fibronectin:CD66b | 38.063 | 20.531 | 57.106 | <0.001 |
| | | CD66b:CD14 | -39.675 | -61.799 | -22.303 | <0.001 |
| | | CD68:CTLA4 | 48.951 | 26.883 | 70.672 | <0.001 |
| | | GZMB:FOXP3 | -23.341 | -42.375 | -6.497 | 0.006 |
| | | CD11c:CD68 | -13.852 | -30.658 | -0.411 | 0.012 |
| | | Histone-H3:S6 | 24.454 | 5.092 | 47.301 | 0.018 |
| | | Histone-H3:CD4 | -18.759 | -38.146 | -1.102 | 0.038 |
| | Away | CD14 | -43.76 | -69.475 | -20.21 | <0.001 |
| | | MLH1 | 48.795 | 24.395 | 82.058 | <0.001 |
| | | Ki-67 | -47.707 | -75.653 | -22.415 | <0.001 |
| | | GAPDH:SMA | 36.206 | 14.351 | 60.277 | <0.001 |
| | | GZMB:CD66b | 44.869 | 25.585 | 68.906 | <0.001 |
| | | CD66b:FAP-alpha | -26.803 | -48.861 | -7.27 | 0.001 |
| | | GZMB | 24.699 | 3.826 | 46.529 | 0.006 |
| | | CD127:ICOS | -18.908 | -42.429 | -0.399 | 0.034 |
| | | GAPDH | 25.898 | 0.963 | 55.777 | 0.05 |
| | | ICOS | -22.103 | -48.788 | 0.183 | 0.056 |
| | | CD45:MLH1 | 17.479 | -0.791 | 37.581 | 0.057 |

### Differential Co-Expression Supplementary Figures and Tables

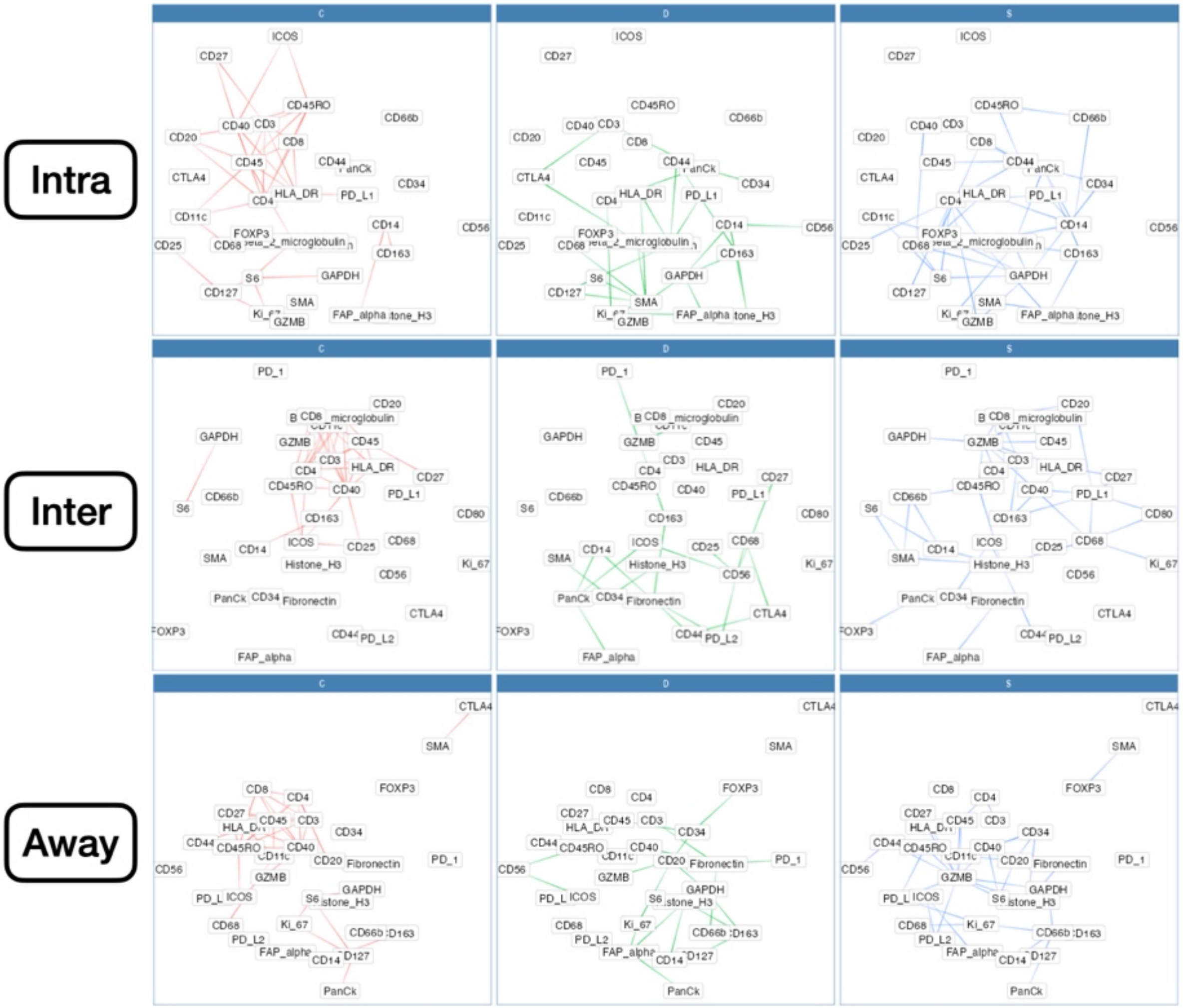


**Supplementary Figure 11: Differential Co-Expression Networks predictive of any metastasis,** stratified by three tissue architectures (*intra, inter, away*). C indicates whether co-expression was conserved between patients with and without metastasis; D indicates whether co-expression differed between patients with and without metastasis; S indicates whether significant co-expression was specific to either patients with or without metastasis. Edges between markers indicate whether relationship could be characterized by C, D or S

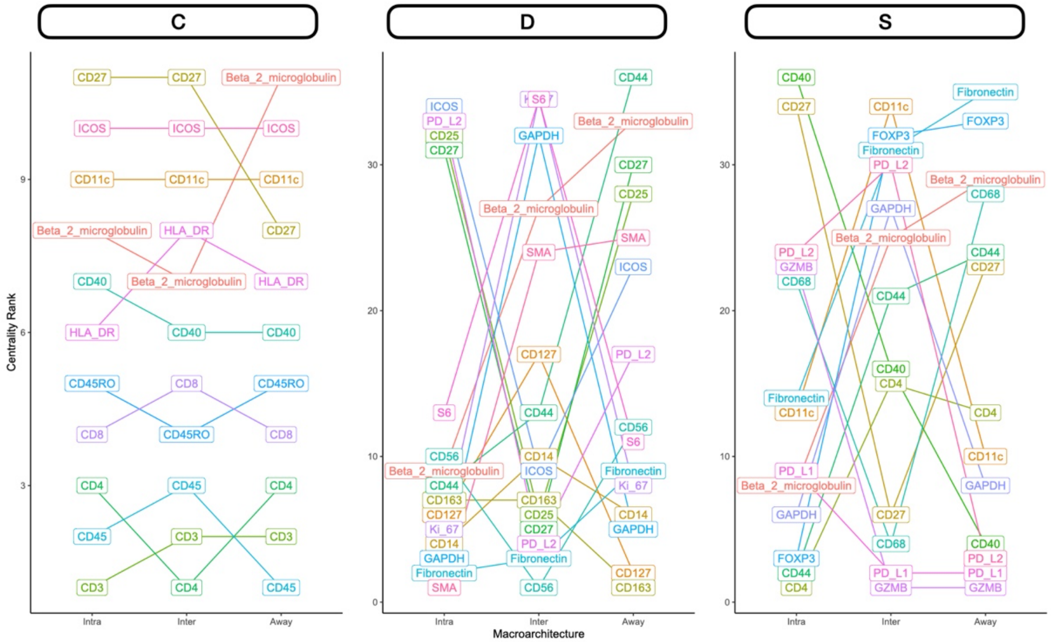


**Supplementary Figure 12: Rank-based summaries of proteins important to the differential co-expression networks predictive of any metastasis,** stratified by three tissue architectures (*intra, inter, away*). C indicates whether co-expression was conserved between patients with and without metastasis; D indicates whether co-expression differed between patients with and without metastasis; S indicates whether significant co-expression was specific to either patients with or without metastasis. Rank indicates eigenvector centrality of protein within each of the networks (lower rank indicates importance in network); proteins with top-10 overall rank were selected for viewing

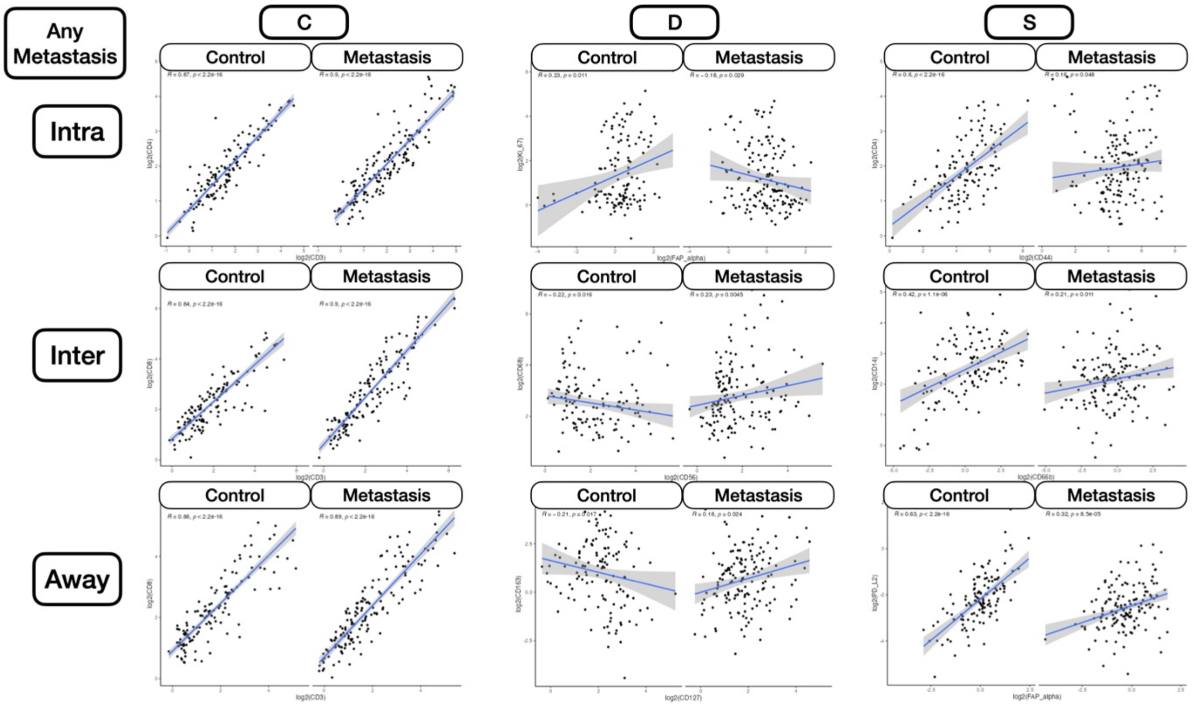

**Supplementary Figure 13: Differential Co-Expression scatterplots between select protein markers, stratified by any metastasis status** within three tissue architectures (*intra, inter, away*). C indicates whether co-expression was conserved between patients with and without metastasis; D indicates whether co-expression differed between patients with and without metastasis; S indicates whether significant co-expression was specific to either patients with or without metastasis

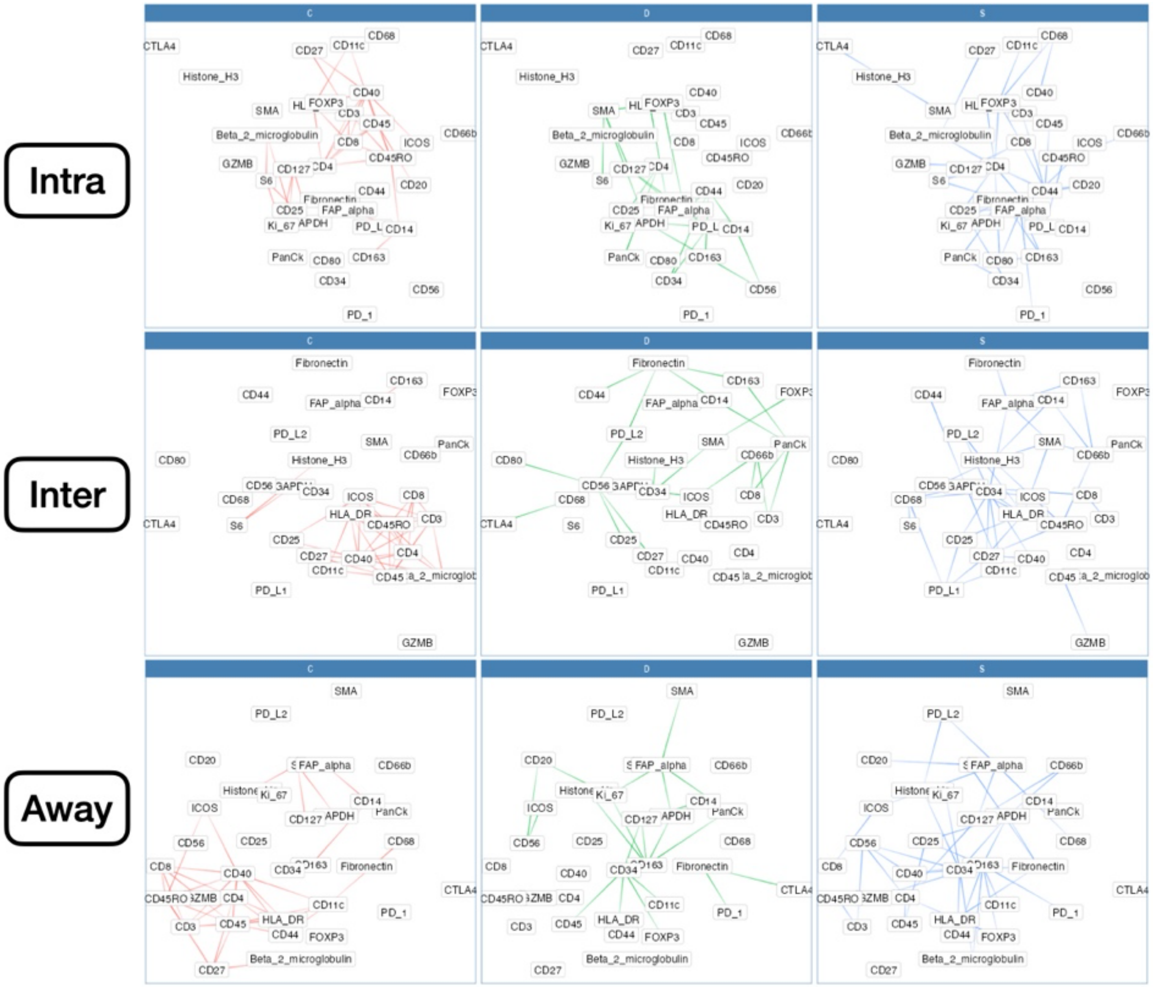


**Supplementary Figure 14: Differential Co-Expression Networks predictive of nodal metastasis,** stratified by three tissue architectures (*intra, inter, away*). C indicates whether co-expression was conserved between patients with and without metastasis; D indicates whether co-expression differed between patients with and without metastasis; S indicates whether significant co-expression was specific to either patients with or without metastasis. Edges between markers indicate whether relationship could be characterized by C, D or S

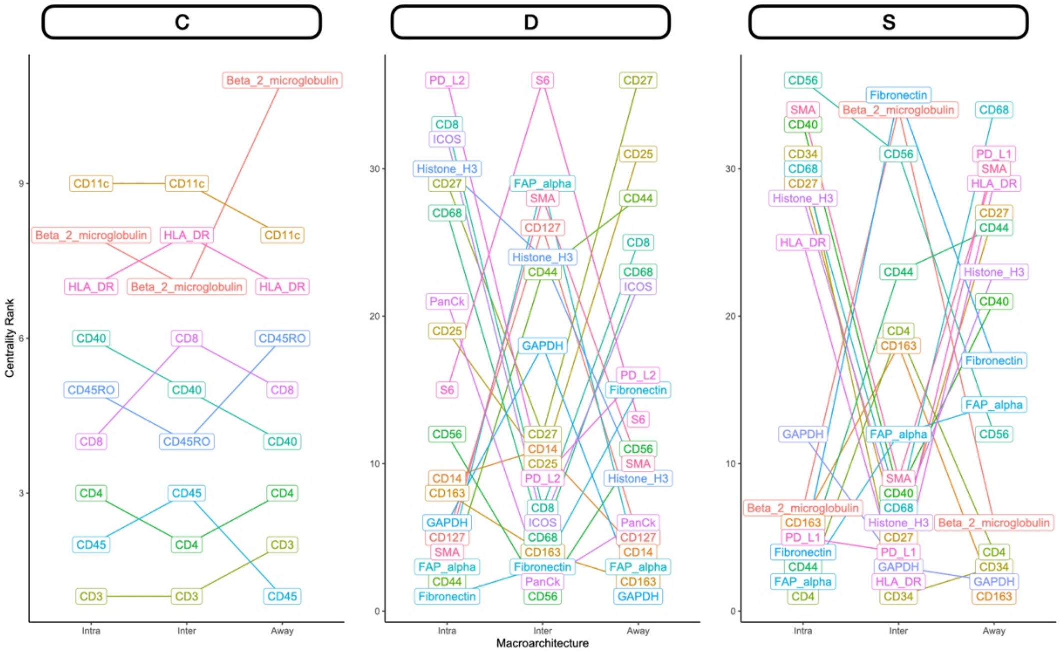


**Supplementary Figure 15: Rank-based summaries of proteins important to the differential co-expression networks predictive of nodal metastasis,** stratified by three tissue architectures (*intra, inter, away*). C indicates whether co-expression was conserved between patients with and without metastasis; D indicates whether co-expression differed between patients with and without metastasis; S indicates whether significant co-expression was specific to either patients with or without metastasis. Rank indicates eigenvector centrality of protein within each of the networks (lower rank indicates importance in network); proteins with top-10 overall rank were selected for viewing

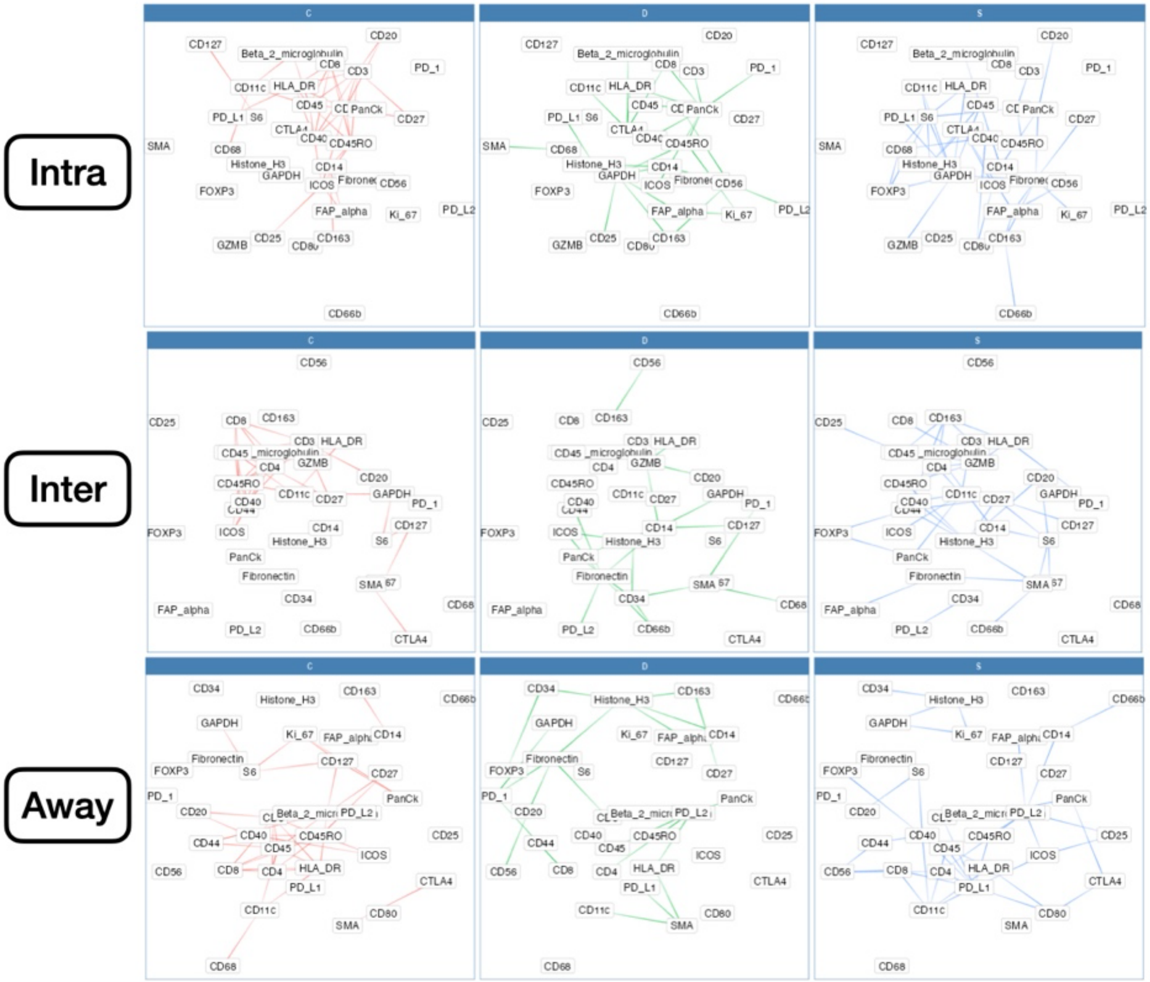


**Supplementary Figure 16: Differential Co-Expression Networks predictive of distant metastasis,** stratified by three tissue architectures (*intra, inter, away*). C indicates whether co-expression was conserved between patients with and without metastasis; D indicates whether co-expression differed between patients with and without metastasis; S indicates whether significant co-expression was specific to either patients with or without metastasis. Edges between markers indicate whether relationship could be characterized by C, D or S

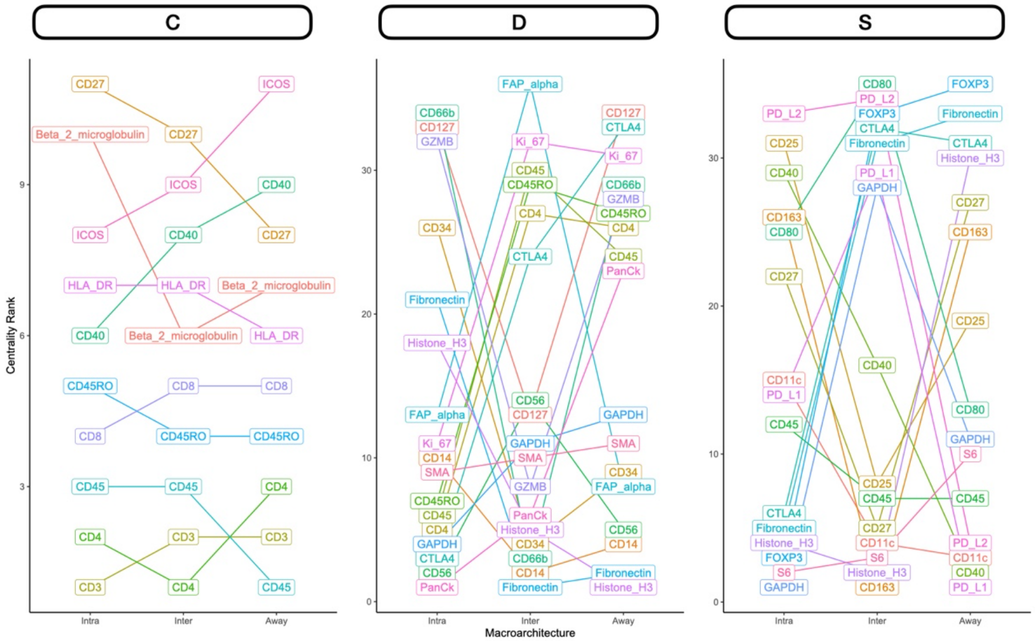


**Supplementary Figure 17: Rank-based summaries of proteins important to the differential co-expression networks predictive of distant metastasis,** stratified by three tissue architectures (*intra, inter, away*). C indicates whether co-expression was conserved between patients with and without metastasis; D indicates whether co-expression differed between patients with and without metastasis; S indicates whether significant co-expression was specific to either patients with or without metastasis. Rank indicates eigenvector centrality of protein within each of the networks (lower rank indicates importance in network); proteins with top-10 overall rank were selected for viewing

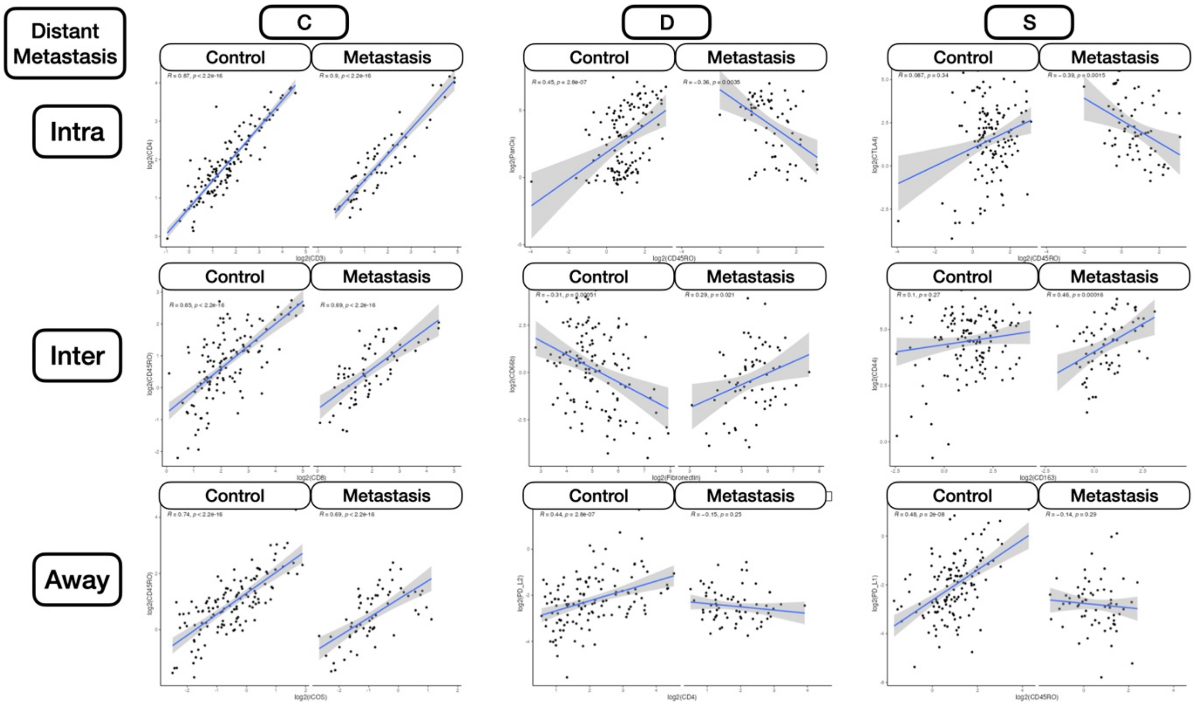

**Supplementary Figure 18: Differential Co-Expression scatterplots between select protein markers, stratified by distant metastasis status** within three tissue architectures (*intra, inter, away*). C indicates whether co-expression was conserved between patients with and without metastasis; D indicates whether co-expression differed between patients with and without metastasis; S indicates whether significant co-expression was specific to either patients with or without metastasis

### Clustering

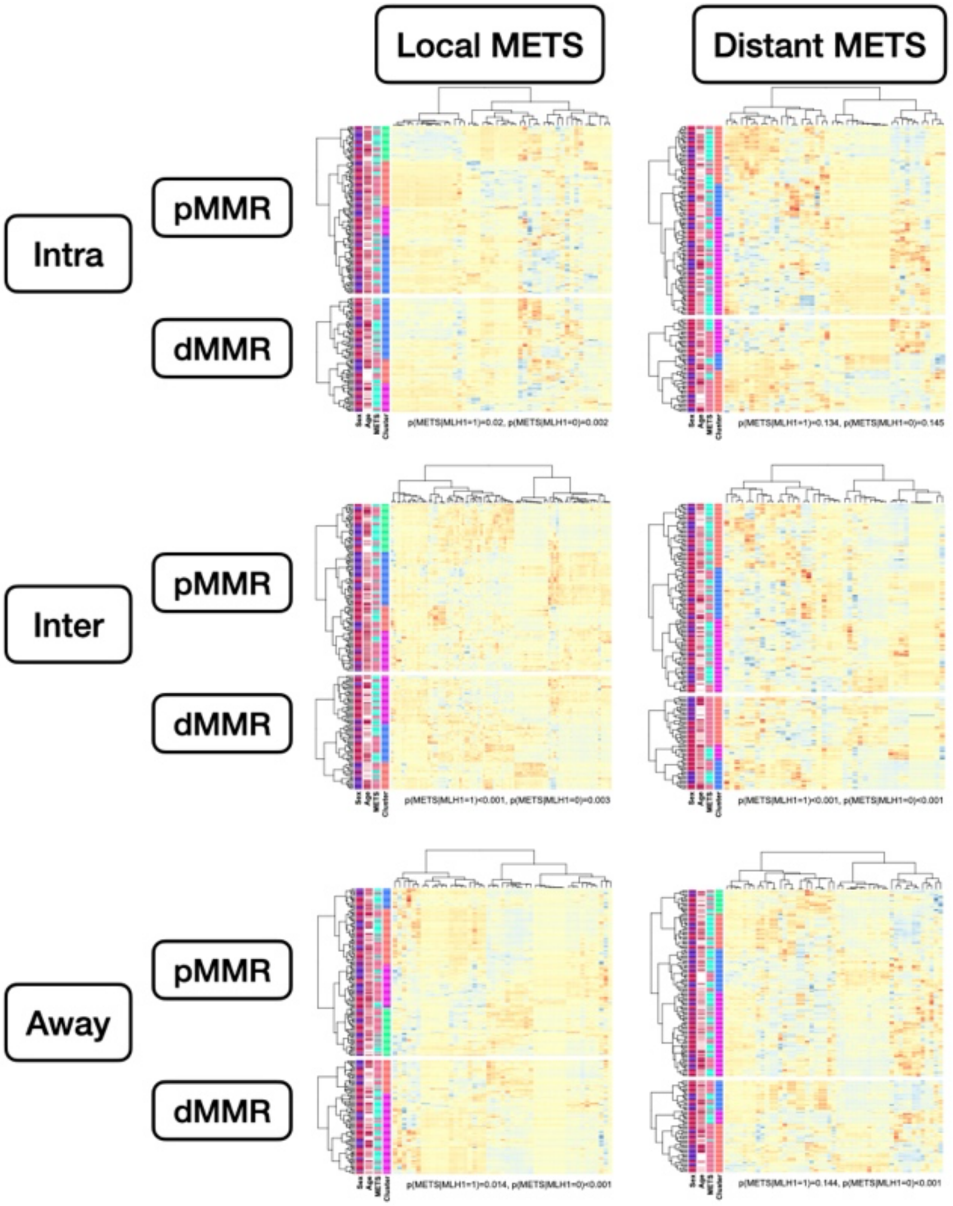


**Supplementary Figure 19: Hierarchical clustering of predictive markers of metastasis extracted from study,** within three tissue architectures (*intra, inter, away*), stratified by MMR-status; fisher's exact tests were used to compare cluster assignment with presence of metastasis to report p-values at bottom of plot to indicate overall associations
